# A unified framework for local-ancestry-aware genetic association analysis across biobanks

**DOI:** 10.64898/2026.08.09.26360047

**Authors:** Linfeng Hu, Taotao Tan, Kai Yuan, Ying Wang, Bram L. Gorissen, Yi-Sian Lin, Pragati Kore, Wenhan Lu, Ravi Mandla, Zhuozheng Shi, Kangcheng Hou, Konrad J. Karczewski, Hailiang Huang, Benjamin M. Neale, Mark J. Daly, Alicia R. Martin, Bogdan Pasaniuc, Elizabeth G. Atkinson, Wei Zhou

## Abstract

Biobanks increasingly include individuals with admixed genomes, yet conventional genome-wide association study frameworks either exclude participants who cannot be confidently assigned to a discrete ancestry group or ignore ancestry-specific effects. We present FELIX, a scalable framework for local-ancestry-aware genetic analysis that retains all participants without requiring discrete ancestry assignment. FELIX combines a compact ancestry-resolved genotype representation (FELIXla) with an adaptive association test that jointly evaluates shared-effect and ancestry-specific models at each variant (FELIXassoc). Simulations demonstrated well-calibrated inference under case-control imbalance and power that adapted to the locus-optimal model. Across 24 phenotypes in 240,038 All of Us participants, FELIX analyzed the 12.1% of individuals excluded by global-ancestry clustering and identified 15.4% more genome-wide significant loci than global-ancestry meta-analysis. Additional discoveries arose from recovering ancestry-specific haplotypes carried by admixed participants and from detecting ancestry-dependent marginal effects. Full-cohort effect estimates also improved polygenic score prediction across ancestries and traits.

## INTRODUCTION

Population biobanks have transformed genetic discovery for complex human traits and diseases by increasing both sample size and ancestral diversity^1^. Individuals with admixed genomes, whose chromosomes comprise segments inherited from multiple ancestral populations, are increasingly represented in biobanks such as the All of Us Research Program (AoU)^2^ and the Million Veteran Program^3^. These cohorts provide important opportunities to broaden genetic discovery across diverse backgrounds. However, commonly used genome-wide association study (GWAS) frameworks remain poorly suited to the mosaic structure of admixed genomes, limiting both statistical power and ancestry-resolved inference.

Two strategies are commonly used to study associations in ancestrally diverse biobanks. Global-ancestry meta-analysis assigns participants to discrete ancestry groups using principal-component or clustering-based approaches, conducts association analyses within each group, and then combines results across groups, as exemplified by the All-by-All framework^4^. This approach can accommodate between-group differences but excludes individuals who cannot be confidently assigned to a single cluster. In AoU global-ancestry meta-analysis^4^, 12.1% of participants fall below the confident threshold for assignment to any global ancestry group, predominantly along African-European-Indigenous American ancestry continua. Mega-analysis instead retains participants in a pooled analysis while adjusting for global ancestry using principal components or related covariates^5^. Although this approach retains more samples, it assumes a shared marginal genetic effect across ancestral backgrounds. Existing approaches therefore require a trade-off between full-cohort analysis and ancestry-specific resolution^5–7^.

Local ancestry inference (LAI) addresses this trade-off by resolving the ancestral origin of chromosomal segments^8^. Methods such as Tractor^9^ and Tractor-Mix^10^ estimate local-ancestry-specific genetic effects using ancestry-resolved allele dosages. These models are advantageous when marginal effects differ across ancestries. But estimation using a multi-degree-of-freedom heterogeneous-effect model throughout the genome loses power relative to a simpler shared-effect test when effects are similar^9,11^, as is expected at many loci^12^. Conversely, traditional shared-effect models can be suboptimal when ancestry-dependent marginal associations arise from differences in allele frequency and linkage disequilibrium^9,13^. The optimal model therefore depends on the locus, yet existing methods generally apply a single effect structure genome-wide. Biobank-scale applications further introduce challenges including computational scalability, relatedness correction, and calibrated inference for highly unbalanced binary traits.

Here we present FELIX (Full-cohort Efficient Local ancestry-Integrated miXed-model framework), a unified framework for biobank-scale local-ancestry-aware genetic analysis that jointly addresses sample retention, local ancestry resolution, and locus-specific genetic architecture, which can vary within an individual’s genome (**Figure 1**). FELIX represents ancestry-resolved allele dosages in FELIXla, a compact binary format designed for efficient storage and variant-level access. The association module, FELIXassoc, evaluates both a shared-effect model based on aggregate dosage and an ancestry-specific model based on local-ancestry-resolved dosages. FELIXassoc then combines evidence from both models using the Cauchy combination test^14^, allowing the association test to approach the power of either model, whether a common effect or ancestry-dependent marginal effect better describes the locus. By modeling ancestry at haplotype resolution within a mixed-model framework, FELIX enables full-cohort association testing while allowing the effect structure to vary across loci.

**Figure 1.**
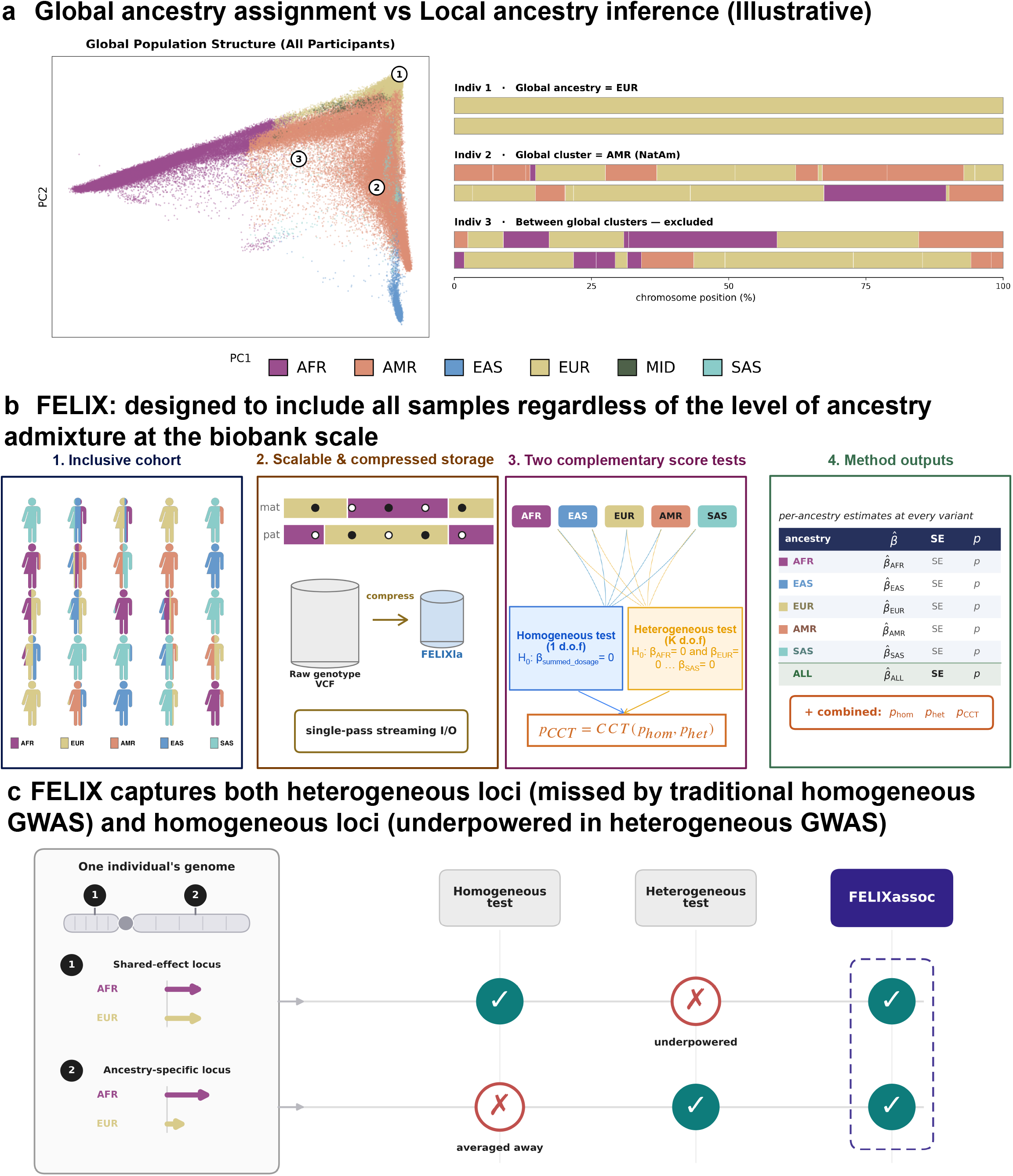
FELIX enables full-cohort, local-ancestry-aware association testing at biobank scale. **a**, Global-ancestry assignment labels each participant with a single ancestry, whereas local-ancestry inference resolves each genome into per-ancestry haplotype segments, motivating full-cohort and finer-scale analysis. **b**, The FELIX workflow. (1) All participants are retained regardless of admixture level. (2) Phased genotypes and inferred local ancestry are stored in FELIXla, a compact, streamable haplotype-resolved format. (3) From a single fitted null model, a homogeneous (shared-effect, 1 d.o.f.) and a heterogeneous (ancestry-specific, *K* d.o.f.) score test are computed and combined by the Cauchy combination test (CCT). (4) FELIXassoc returns, per variant, the per-ancestry effect estimate 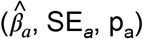, an aggregate ancALL estimate equal to the homogeneous result (i.e., the standard global-ancestry GWAS output), the combined *p*_CCT_, and per-ancestry allele frequencies and local-ancestry haplotype counts. **c**, Because power tracks the better-fitting model at each locus, FELIXassoc detects both shared-effect loci (underpowered in a heterogeneous-only test) and ancestry-specific loci (averaged away in a homogeneous-only test). *K*, number of local ancestries; d.o.f., degrees of freedom; AFR, African; EUR, European; EAS, East Asian; SAS, South Asian; MID, Middle Eastern; AMR, American (Admixed American at global cluster level and Indigenous American at haplotype level). Panels are illustrative schematics. Panel b1 was created using elements from NIAID Visual and Medical Arts, NIH BioArt Source (https://bioart.niaid.nih.gov/bioart/13).

We evaluated FELIX through simulations and analyses of 240,038 AoU^2^ participants using five-way local ancestry inference. FELIXassoc maintained calibration under multi-way admixture and severe case-control imbalance. Simultaneously, FELIXassoc achieved power close to the better-performing shared-effect or ancestry-specific model across genetic architectures. Applied to 24 AoU phenotypes at 1,042,379 HapMap3^15^ markers, FELIXassoc retained the 12.1% of participants excluded by global ancestry classification and identified additional signals through two complementary mechanisms: recovery of ancestry-specific haplotypes carried by admixed individuals and detection of ancestry-dependent marginal effects obscured by shared-effect models. Full-cohort effect estimates from FELIX also improved polygenic score prediction across four independent UK Biobank^16^ validation ancestries, outperforming both global-ancestry meta-analysis and global-ancestry-matching training. Together, these results establish FELIX as a scalable framework for full-cohort analysis at biobank-scale while producing accurate, ancestry-resolved summary statistics for downstream genetic analyses.

## RESULTS

### Overview of the FELIX framework

FELIX provides a unified framework for local-ancestry-aware association testing in biobank-scale cohorts (**Figure 1, Supplementary Figure 1, Online Methods**). Following local ancestry inference (LAI), genotypes are decomposed into ancestry-specific alternative-allele dosages and local ancestry haplotype counts for each variant and individual. These data are stored in FELIXla, a compact binary format that supports efficient variant-level access without repeatedly reconstructing ancestry-resolved dosages from phased genotype files. The FELIXla representation serves as efficient input to FELIXassoc at the biobank scale.

FELIXassoc performs association testing in two steps. First, it fits a generalized linear mixed model under the null hypothesis of no genetic effect, estimating fixed effects and variance components while accounting for covariates, population structure, and sample relatedness through an ancestry-adjusted sparse GRM^17^. This null model is fitted once and reused for all variants and association models leveraging the SAIGE framework^18^. Second, FELIXassoc evaluates two complementary tests at each variant. A one-degree-of-freedom shared-effect test uses the aggregate alternative-allele dosage across ancestral backgrounds, whereas a K-degree-of-freedom ancestry-specific test jointly evaluates the K local-ancestry-resolved dosages. For binary traits, Saddlepoint approximation^19^ is applied to the marginal score statistics to maintain calibration under case-control imbalance. Evidence from the shared-effect and ancestry-specific tests is then combined using the Cauchy combination test^14^, allowing FELIXassoc to adapt to locus-specific genetic architecture without requiring one association model to be selected genome-wide.

At loci where local ancestry itself is associated with the phenotype, FELIXassoc can additionally condition each ancestry-specific dosage on the corresponding local ancestry haplotype count. This option helps distinguish allelic associations from associations driven by local ancestry and admixture linkage disequilibrium and can improve the interpretation of ancestry-specific effect estimates.

### FELIXassoc maintains calibration under multi-way admixture and case-control imbalance

FELIXassoc maintained well-controlled type I error across multi-way admixture, low-frequency variation and severe case-control imbalance (**Supplementary Figure 2**). We simulated a three-way African-European-Indigenous American admixed cohort (mean ancestry proportions of approximately 10% African (AFR), 55% European (EUR) and 35% Indigenous American (AMR) to broadly resemble the composition of admixed Americans). The simulated cohort included 10,000 individuals, half unrelated and half from 500 three-generation families. Ancestry-specific allele frequencies were generated using a three-population Balding-Nichols model^20^ (**Online Methods**). We evaluated type I error for low-frequency variants (minor allele frequencies of 1-5%) and common variants (MAF greater than 5%). Simulations included quantitative traits and binary traits with prevalence of 10% and 1%.

Across 10 million independent null simulations, the shared-effect (test under the assumption of homogeneous effects across ancestries, denoted as *test*_*hom*_), ancestry-specific (test under the assumption of heterogeneous effects across ancestries, denoted as *test*_*het*_), and Cauchy-combined (*test*_*CCT*_) tests, as well as ancestry-specific tests all maintained well-controlled type I error (**Supplementary Figure 2**). Calibration was preserved for low-frequency variants at binary traits with 1% prevalence, indicating that Saddlepoint approximation^19^ remained effective when applied to sparse local-ancestry-resolved dosages. Under the same simulation settings, Tractor-Mix^10^ remained well-calibrated for quantitative traits and for binary traits of 10% prevalence but showed inflation at 1% prevalence in both the joint and ancestry-specific summary statistics (**Supplementary Figure 3**). These results demonstrate that FELIXassoc maintains calibrated association testing across variant-frequency spectra and trait types, including severe case-control imbalance under multi-way admixture, extending local-ancestry-aware methods to low-prevalence traits.

### FELIXassoc adapts to shared and ancestry-specific genetic architectures

FELIXassoc maintained high statistical power across loci with different cross-ancestry genetic architectures by combining complementary shared-effect and ancestry-specific tests. We evaluated power across 100 simulated phenotypes under two representative architectures: a homogeneous architecture in which the causal effect was shared across the three ancestral backgrounds (*β*_*AFR*_= *β*_*EUR*_ = *β*_*AMR*_) and an asymmetric heterogeneous architecture in which the effects differed among ancestries (*β*_*AFR*_, *β*_*EUR*_, *β*_*AMR*_) = (0, 0.5*β, β*)). Power was defined as the proportion of simulations in which the causal variant reached genome-wide significance.

Under the homogeneous architecture, the one-degree-of-freedom shared-effect test (test_hom_) was most powerful as the simulated causal effect was homogeneous across ancestries, whereas the ancestry-specific test (test_het_) incurred a degrees-of-freedom penalty by estimating multiple effects when a single parameter was sufficient. Under the heterogeneous architecture, test_het_ substantially outperformed test_hom_, recovering signals that were attenuated when effects were pooled across ancestral backgrounds.

Across the evaluated settings, the Cauchy-combined test (test_CCT_) closely tracked the power of the better-performing component test (**Supplementary Figure 4**). FELIXassoc therefore remains sensitive to both shared and ancestry-dependent marginal effects without requiring the effect structure to be specified in advance. This flexibility becomes increasingly important as the number of modeled ancestral backgrounds increases, because the degrees-of-freedom penalty of an ancestry-specific test grows with K, making a uniformly heterogeneous model progressively less efficient when effects are shared. Although this penalty was modest in the three-way simulations, it is expected to be more pronounced in multi-way analyses such as the five-ancestry AoU^2^.

### FELIXassoc scales to biobank-sized cohorts

FELIXassoc evaluates shared-effect and ancestry-specific association models using a single fitted null model, enabling local-ancestry-aware testing with computational costs comparable to conventional global-ancestry-based mixed-model association analysis (**Supplementary Table 1**). In benchmarks testing 1,000 variants under a 20-hour wall-time limit, Tractor-Mix exceeded available memory when the sample size surpassed 50,000 individuals, whereas FELIXassoc continued to scale to larger cohorts (**Supplementary Table 2**). For a representative AoU phenotype (*N*_*case*_ = 5,007, *N*_*control*_ = 185,284), FELIXassoc analyzed 1,042,379 HapMap3^15^ variants in 36.8 CPU-hours, using 4.54□GB of peak memory.

Three design features enable this scalability (**Online Methods**). First, FELIXassoc retrieves ancestry-specific dosages directly from the FELIXla representation rather than reconstructing them from phased genotypes at each variant. Second, it inherits the computational structure of SAIGE^18^: variance components are estimated once under the null model, and a variance-ratio approximation allows variant-level score statistics to reuse precomputed^21–23^, covariate-projected quantities rather than requiring repeated matrix operations. Third, the shared-effect and ancestry-specific tests are computed from the same fitted null model, and the Cauchy combination^14^ requires only a closed-form calculation. FELIXassoc can therefore evaluate complementary genetic architectures without fitting separate mixed models for each test.

### FELIX enables full-cohort genetic association analysis in All of Us

We applied FELIXassoc to 240,038 participants from the All of Us Research Program^2^ (v7), analyzing the full cohort without exclusions based on global ancestry assignment (**Supplementary Figure 5**). Local ancestry calls were available for HapMap3^15^ variants and decomposed each haplotype into African (AFR), European (EUR), East Asian (EAS), South Asian (SAS), or Indigenous American ancestry (AMR).

By operating on local ancestry rather than assigning each participant to a single global ancestry group, FELIXassoc retained the 12.1% of AoU participants (N=25,821) excluded from the All-by-All global-ancestry meta-analysis^4^ because they did not meet the posterior-probability threshold for assignment to any of their discrete global ancestry clusters. Among these participants, the AoU classifier’s highest-probability ancestry category was European for 63.7%, African for 18.9%, admixed American for 14.6%, East Asian for 1.0%, South Asian for 0.8%, and Middle Eastern for 0.8%. In principal component space, these participants were concentrated along the African-European-admixed American ancestry continua, consistent with recent admixture rather than membership in a single continental ancestry cluster^24^ (**Figure 2a-d**).

**Figure 2.**
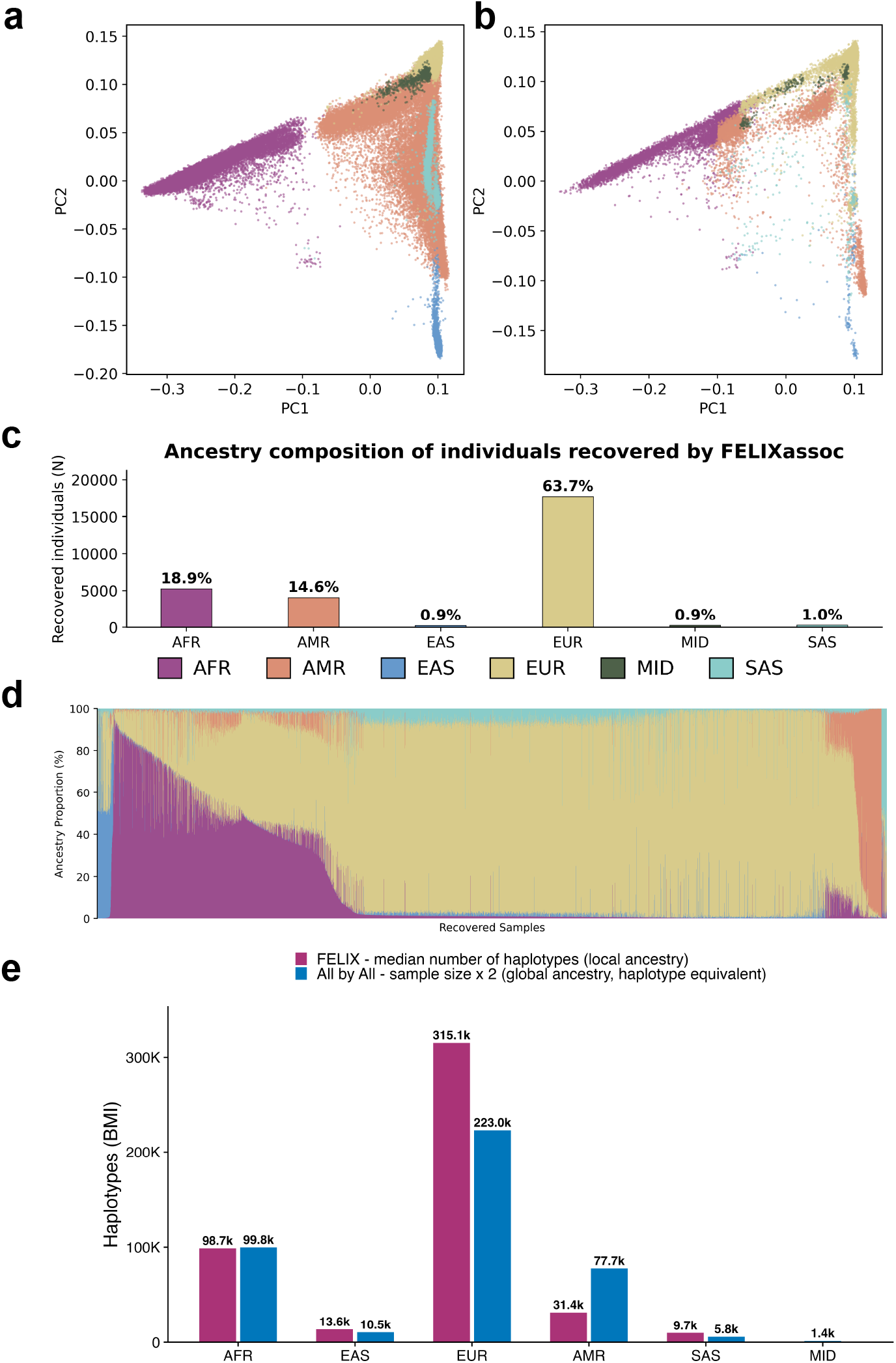
Full-cohort inclusion in the All of Us Research Program. **a-d**, Genetic ancestry of All of Us participants in projected principal-component space (reference PCA space defined by the Human Genome Diversity Project and 1000 Genomes Project reference panel, gnomAD v3.1.2). **a**, Participants included in the All-by-All global-ancestry meta-analysis. **b**, The same PC1-PC2 space showing the participants recovered by FELIX (12.1% of the cohort) that global-ancestry analysis excluded because they fell below the ancestry classifier confidence threshold. **c**, Ancestry composition of the recovered participants. **d**, Admixture proportions of representative highly admixed recovered participants. **e**, All of Us v7 effective sample size, expressed as haplotype counts, under global-ancestry versus local-ancestry analysis for an example trait (body mass index, as this physical measurement is recorded for the majority of participants). Median N_haplo_ refers to the median number of haplotypes across the entire genome examined. Notation: AFR, African; EAS, East Asian; EUR, European; AMR, Indigenous American; SAS, South Asian; MID, Middle Eastern.

We analyzed 24 quantitative and binary phenotypes, including height, body mass index (BMI), circulating biomarkers and disease outcomes (**Supplementary Table 3**). Thus, every participant and each ancestry-specific haplotype carried by that participant contributed to the relevant association tests.

### FELIXassoc increases discovery while remaining concordant with global-ancestry meta-analysis

We compared FELIXassoc with the fixed-effect All-by-All global-ancestry meta-analysis performed in the same AoU release (All-by-All v7^4^). Comparisons were restricted to variants tested by both approaches and therefore assess concordance and relative discovery within the shared marker set rather than across all variants available to the All-by-All^4^ analysis. Across the 24 phenotypes, the FELIXassoc shared-effect, ancestry-specific, and Cauchy-combined tests (test_het_, test_hom_, and test_CCT_), as were the tests for each of the five local ancestry components, showed well-calibrated statistics (**Supplementary Figure 6**).

Effect estimates from the FELIXassoc shared-effect test were strongly concordant with those from the global-ancestry meta-analysis (Pearson’s r = 0.96; **Supplementary Figures 7 and 8**), as expected because both approaches model a shared marginal effect across ancestral backgrounds. Within the shared marker set, however, the FELIXassoc combined test identified 837 independent genome-wide significant loci, compared with 725 loci identified by global-ancestry meta-analysis, representing a 15.4% increase. FELIXassoc recovered 679 of the 725 baseline loci (93.7%).

Most method-specific lead signals reflected differences in power near the genome-wide significance threshold or selection of different index variants, rather than disagreement about the presence of an association. Among the 158 FELIXassoc-specific lead signals, 103 (65%) were near-significant (5 × 10^−8^ < *p* < 5 × 10^−6^) in the baseline analysis, 34 (22%) showed weaker evidence, and only 21 (13%) were largely missed. Conversely, 41 of the 46 baseline-specific signals were near-significant in FELIXassoc, and only one was largely missed. In addition, 72.7% of FELIXassoc-specific lead variants were located within 500kb of a genome-wide significant baseline signal represented by a different index variant.

FELIXassoc ancestry-specific test contributed a distinct subset of associations not captured by shared-effect testing, whereas the shared-effect test retained high power at loci with concordant effects. Combining the two tests therefore increased discovery without substantially compromising recovery of associations detected by conventional global-ancestry meta-analysis.

### Full-cohort local-ancestry analysis improves precision at established loci

At the 725 loci significant in the global-ancestry meta-analysis, the FELIXassoc shared-effect analysis reduced the standard error by a median of 8.4% (reduced at 99% of loci) and raised the chi-squared statistic by a median of 8.6%, with concordant effect directions (**Supplementary Tables 4 and 5**). The precision gain reflected the ancestry of participants recovered by the full-cohort analysis. Relative to the global ancestry strata, ancestry-specific standard errors decreased by 19% (EUR), 20% (EAS) and 46% (SAS), whose haplotypes were often carried by individuals excluded from discrete global groups. The reduction was 9% for the already well-represented African group.

The comparison differed for Indigenous American ancestry. The admixed American global ancestry group (up to N=38,971) combines Indigenous American, European, and African haplotypes and is therefore larger than the Indigenous American local ancestry segments (up to N ≈ 15,738). Because local ancestry was inferred against a more specific Indigenous American reference than AoU global-ancestry inference (**Online Methods**), FELIXassoc trades nominal sample size for higher specificity on Indigenous American haplotypes, while non-matching haplotypes are reassigned to other backgrounds, predominantly European. The precision gain therefore reflects both increased effect sample size and more accurate allocation of haplotypes.

### Recovering ancestry-specific haplotypes carried by admixed participants increases association power

The first mechanism underlying additional discovery was increased effective sample size through the recovery of ancestry-specific haplotypes carried by admixed individuals. Global ancestry analysis assigns the entirety of each individual genome to one ancestry stratum or excludes the individual altogether. FELIXassoc instead assigns each haplotype segment to its inferred local ancestry, allowing ancestry-specific alleles carried by every participant to contribute to the corresponding test.

The established *IL23R R381Q* coding variant (rs11209026), which protects against Crohn’s disease^25,26^, illustrates this gain. The association reached genome-wide significance with FELIXassoc (*p*_*cct*_ = 5.8 × 10^-9^), compared with (*p* = 7.7 × 10^-7^) in the global-ancestry meta-analysis, a roughly 130-fold difference in statistical significance (**Figure 3a, Supplementary Figure 9** and **Supplementary Table 6**). The signal was driven primarily by EUR haplotypes (*β*_*EUR*_ = -0.46, *p =4*.9 × 10^-9^). Because the estimated EUR allele frequency was similar under the local and global ancestry classifications, the gain was attributable primarily to increased effective sample size. Incorporating European haplotypes carried by admixed participants increased the effective European sample size by 43.2%, from 99,252 to 142,174, reducing the standard error sufficiently for the association to become significant.

**Figure 3.**
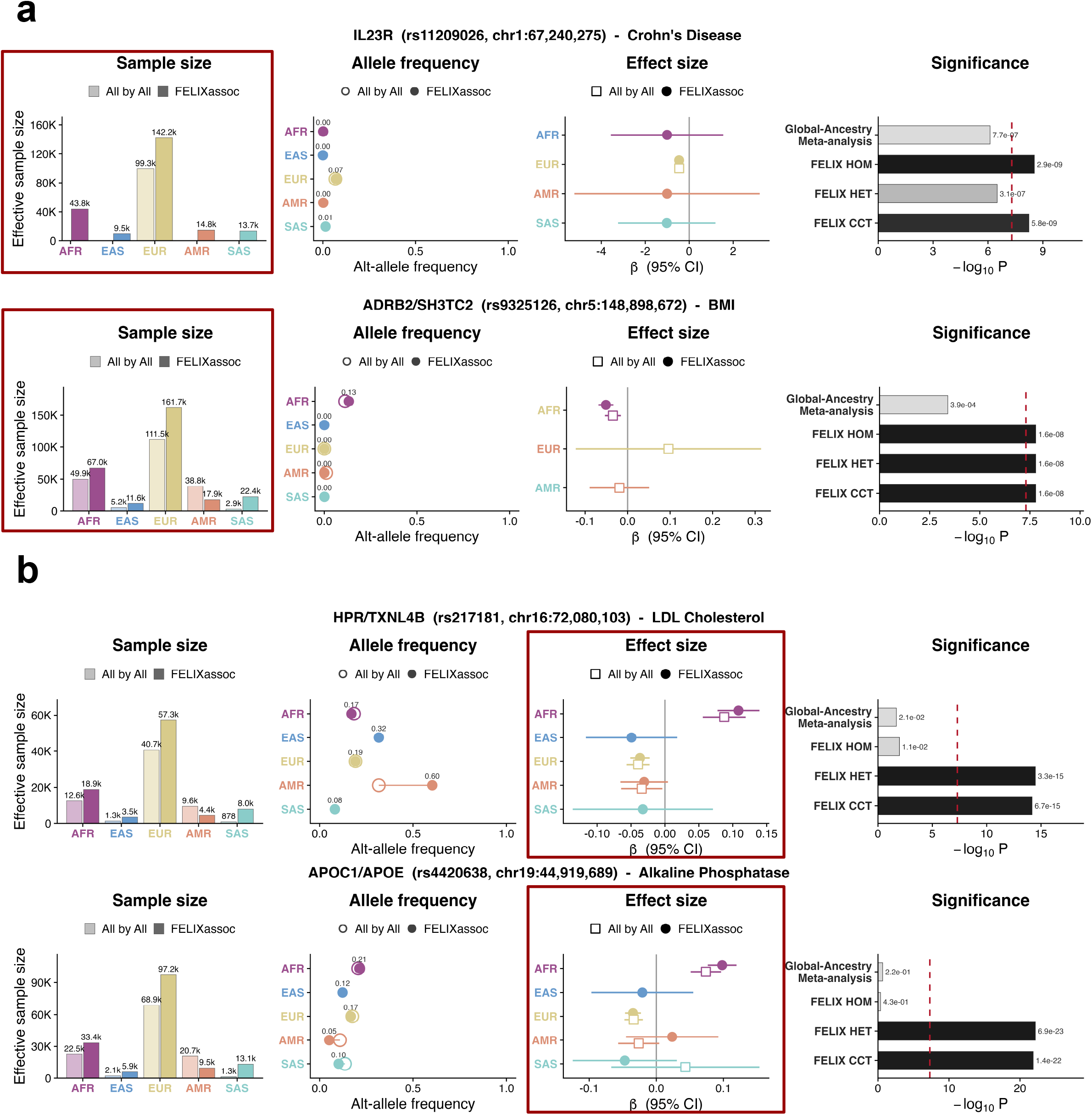
FELIXassoc discovery mechanisms at example loci. **a**, Recovery of under-represented ancestry-specific haplotypes carried by admixed participants increases the effective sample size and statistical power. **b**, Modeling ancestry-specific marginal effects recovers cross-ancestry associations that are attenuated by effect cancellation under a shared-effect model.

The same mechanism also increased power for non-European ancestry signals. At rs9325126 in the *ADRB2-SH3TC2* region, an AFR association with BMI reached significance in FELIXassoc (p_CCT_ = 1.6×10^-8^) but not in the global-ancestry meta-analysis (p = 3.9×10^-4^). The proportion of participants contributing AFR haplotypes at this locus increased from 11% under global assignment to 13% under local ancestry analysis, providing additional effective sample size for the ancestry-specific association.

Local ancestry resolution also yields allele-frequency estimates that were more specific to the relevant ancestral background. At the *PNPLA3 I148M* region, a well-established determinant of hepatic fat accumulation and liver transaminase levels^27–29^, FELIXassoc estimated an AMR ancestry allele frequency of 0.79, consistent with previous estimates in Indigenous American populations^30^. But the admixed American global ancestry group had an allele frequency of 0.44 because it combined Indigenous American, European, and African haplotypes. Resolving the variant by local ancestry produced a more accurately scaled Indigenous American effect.

Together, these examples show that haplotypes carried by admixed individuals can increase ancestry-specific effective sample sizes, refine allele-frequency estimates and strengthen associations that remain below genome-wide significance in analyses based on discrete global ancestry groups.

### Modeling ancestry-specific effects recovers associations obscured by effect cancellation

A second mechanism of discovery arose when marginal effects differed across ancestral backgrounds. Fixed-effect global-ancestry meta-analysis estimates a common effect and can therefore attenuate associations when ancestry-specific estimates differ substantially or point in opposite directions. The FELIXassoc ancestry-specific test retains such signals, while test_CCT_ preserves sensitivity to loci with either shared or heterogeneous effects.

At the *APOE-APOC1* locus, rs4420638 showed ancestry-dependent associations with alkaline phosphatase levels. The global-ancestry analyses estimated significant effects in opposite directions in the African and European strata, but their fixed-effect combination was null (p=0.22). Consistent with this result, the FELIXassoc shared-effect test was also null (p=0.43).

In contrast, FELIXassoc estimated a positive African-ancestry effect (β_AFR_ = 0.098, p = 2.7×10^-19^) and a negative European-ancestry effect (β_EUR_ = -0.035, p = 9.5×10^-9^). The ancestry-specific test strongly detected the association (p= 6.9×10^-23^), resulting in a combined p-value of 1.4×10^-22^ **(Figure 3b** and **Supplementary Table 8**).

A similar pattern was observed at rs217181 in the *HPR-TXNL4B* region for LDL cholesterol. Opposing estimates in the baseline African (β_AFR_ = 0.087, p = 6×10^-8^) and European (β_EUR_ = -0.04, p = 4.5×10^-6^) global strata results largely canceled in the meta-analysis (p=0.02) and in the FELIXassoc shared-effect test (p=0.01). FELIXassoc recovered the association through the ancestry-specific test (p_het_ = 3.3×10^-15^) (**Figure 3b** and **Supplementary Table 8**). This ancestry-dependent association is consistent with previous evidence that structural and allelic variation at *HPR* differs across continental populations and influences lipid metabolism^31,32^.

These associations demonstrate why neither a uniformly shared-effect nor a uniformly ancestry-specific model is optimal. Shared-effect analyses can lose power through attenuation or cancellation of ancestry-dependent effects, whereas ancestry-specific tests incur a degrees-of-freedom cost when effects are concordant. By evaluating both models and combining their evidence, FELIXassoc remains sensitive across a full range of cross-ancestry marginal effect patterns.

### Full cohort effect estimates improve polygenic score prediction across ancestries

We next examined whether the gains from analyzing the full cohort extended beyond association discovery to polygenic score (PRS) prediction. Using PRS-CS^33^, we constructed scores from AoU summary statistics for the 11 quantitative traits analyzed by FELIXassoc and evaluated prediction in independent European, African, East Asian, and Central/South Asian UK Biobank^16^ cohorts defined using Pan-UK Biobank^34^ ancestry assignments (N = 451,509, 6,637, 2,709, and 8,876, respectively). We compared scores derived from the FELIXassoc full-cohort estimates with those derived from the All-by-All global-ancestry meta-analysis and the corresponding ancestry-matched All-by-All analysis^4^. Prediction was quantified as the incremental R^2^ relative to a baseline model containing 20 genetic principal components, age, sex, and their interactions, with 95% confidence intervals obtained by resampling validation individuals (**Online Methods**).

Scores derived from the FELIXassoc full-cohort estimates achieved the highest overall prediction accuracy in each validation cohort (**Figure 4 and Supplementary Table 9**). In the European cohort, the FELIXassoc score outperformed both the global-ancestry meta-analysis and ancestry-matched scores for all 11 traits (median incremental R^2^ 3.6% versus 3.3% and 1.3%). The largest improvement was observed for alkaline phosphatase, for which incremental R^2^ increased to 12.8%, compared with 11.2% and 10.8% for the two comparator scores. More modest gains were observed for lipid traits and physical measurements.

**Figure 4.**
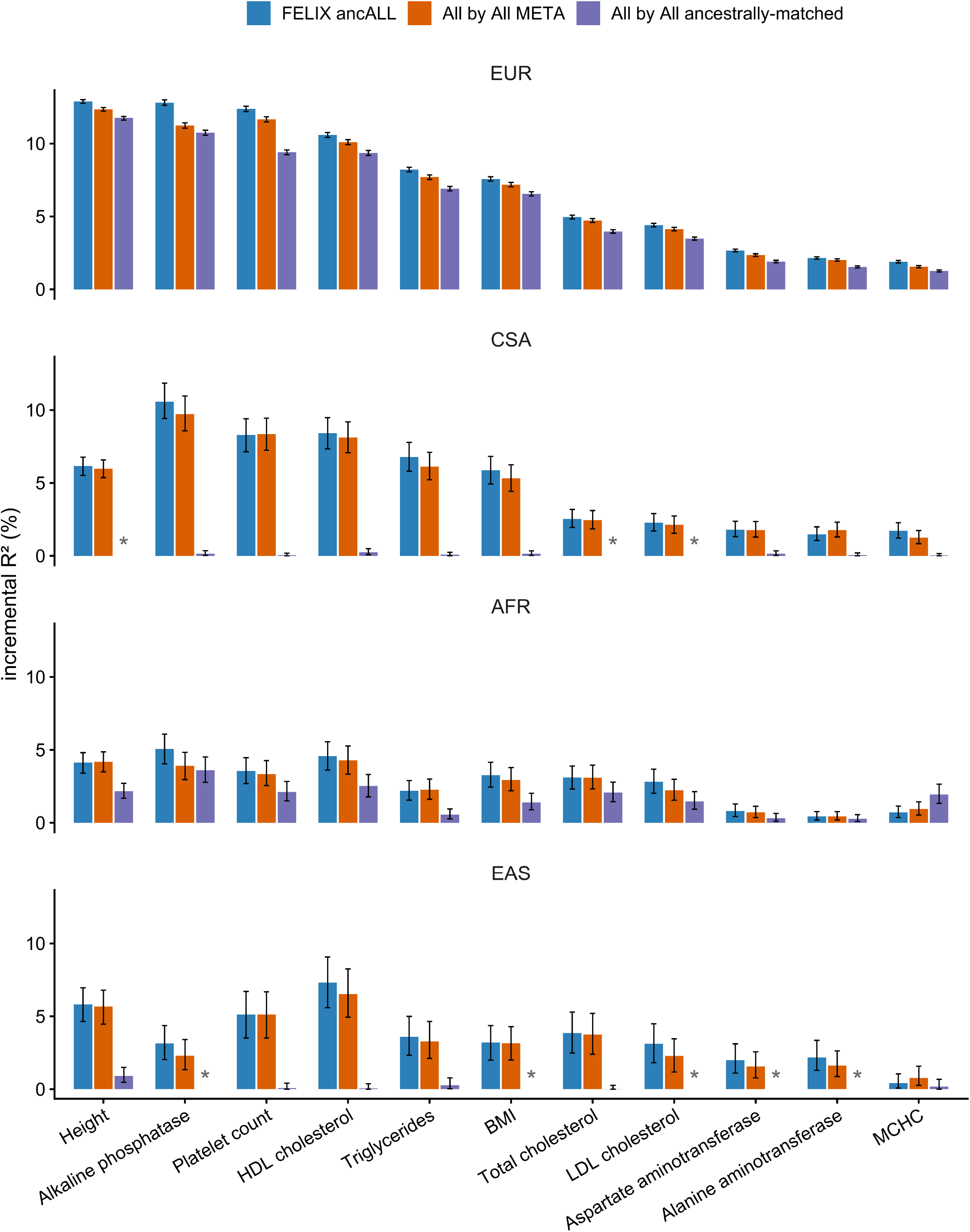
Polygenic prediction in the UK Biobank from All of Us summary statistics. Incremental variance explained (ΔR^2^, %) by three sets of polygenic scores in 4 independent UK Biobank validation cohorts, defined by Pan-UK Biobank ancestry assignment: EUR (European), CSA (Central/South Asian), AFR (African), and EAS (East Asian). Discovery summary statistics were computed in All of Us. For every trait, three predictors are compared: FELIX ancALL, the full-cohort effect estimate from FELIXassoc (all local ancestries analyzed jointly); All by All META, the global-ancestry-based meta-analysis of the All-by-All ancestry-stratified GWAS; and All by All ancestrally-matched, the single All-by-All GWAS from the global ancestry matching each validation ancestry (European, N = 451,509; African, N = 6,637; East Asian, N = 2,709; Central/South Asian, N = 8,876). For each trait and predictor, the bar plot shows the distribution of incremental R^2^ over 2,000 bootstrap resamples of the validation individuals. Ancestries denoted * represent missing GWAS (due to limited sample size in All-by-All per-global-ancestry strata analyses).

The improvement was not restricted to European prediction. Relative to the global-ancestry meta-analysis, the FELIXassoc full cohort score performed better for 8 of 11 traits in the African cohort and for 9 of 11 traits in both the East Asian and Central/South Asian cohorts. It also outperformed the corresponding ancestry-matched score for 10, 6, and 8 traits, respectively. The largest differences occurred where ancestry-matched discovery sample sizes were smallest. In the East Asian and Central/South Asian validation cohorts, scores derived from the corresponding AoU ancestry-stratified analyses explained little phenotypic variance (median incremental R^2^ approximately 0.1%). By contrast, FELIXassoc made use of the haplotype information to achieve median incremental R^2^ values of 3.2% and 5.9%, respectively. Together, these findings demonstrate that the gains from full-cohort, local-ancestry-aware association translate into improved downstream polygenic prediction across ancestries.

## DISCUSSION

We developed FELIX, a unified framework for full-cohort local-ancestry-aware genetic analysis in biobanks, pairing the compact FELIXla genotype representation with FELIXassoc, the association test evaluating shared and ancestry-specific effects at each variant while retaining every participant. By combining both shared-effect and ancestry-specific effect models from a single null-model fit, FELIXassoc attains power tracking the locus-optimal test without imposing one model of cross-ancestry genome effects across the genome. This adaptive strategy is particularly useful in multi-way admixed cohorts, where the degrees-of-freedom penalty of ancestry-specific testing increases with the number of modeled ancestries. The framework also addresses statistical and computational challenges encountered in biobanks. An ancestry-adjusted sparse genetic relationship matrix and Saddlepoint approximation^19^ keep association calibrated across continuous ancestry and under severe case-control imbalance. Reusing a single null-model fit and accessing ancestry-resolved dosages directly through FELIXla enabled practical analysis of hundreds of thousands of participants. Altogether, FELIX provides a common computational and statistical foundation for scalable local-ancestry-aware association discovery and downstream genetic analysis in increasingly diverse biobanks^35^.

Application to the All of Us Research Program^2^ demonstrated that sample retention and ancestry resolution are achieved simultaneously. Global-ancestry meta-analysis^4^ excluded 12.1% of participants who could not be confidently assigned to a discrete ancestry group. FELIXassoc retained these participants and allowed each ancestry-specific haplotype to contribute to the corresponding association test. Within the shared set of variants, FELIXassoc identified 837 loci reaching genome-wide significance, compared with 725 from global-ancestry meta-analysis, while remaining strongly concordant with the baseline results.

Additional discoveries arose through two mechanisms. First, FELIXassoc recovered ancestry-specific haplotypes carried by admixed participants, increasing effective sample size and improving ancestry-specific allele-frequency estimates, as illustrated by *IL23R, ADRB2-SH3TC2* and *PNPLA3*. Second, FELIXassoc detected loci with ancestry-dependent marginal effects that were attenuated under shared-effect models. At *APOE-APOC1* and *HPR-TXNL4B*, opposing African- and European-ancestry effects produced weak evidence in fixed-effect meta-analysis but strong evidence in the ancestry-specific test. Such heterogeneity may reflect differences in linkage disequilibrium, allele frequency, haplotype structure or environmental context rather than ancestry-specific causal biology.

The advantages of full cohort, local-ancestry-aware association from FELIXassoc also extended to downstream polygenic prediction. Across four independent UK Biobank ancestry groups^16,34^, polygenic scores derived from FELIXassoc full-cohort effect estimates consistently outperformed or matched those derived from conventional global-ancestry meta-analysis and ancestry-stratified analyses. These findings suggest that modelling local ancestry while retaining all participants yields summary statistics that are both more precise and more powerful than those obtained by partitioning cohorts into discrete ancestry groups. More broadly, they illustrate that the benefits of local-ancestry-resolved association analysis extend beyond increased locus discovery to improve the quality of effect size estimates for downstream genetic analyses.

Several limitations warrant consideration. First, FELIX inherits uncertainty from local ancestry inference (LAI), and errors in ancestry assignment may attenuate statistical power. LAI accuracy depends on haplotype phasing, reference panel quality and representativeness, marker density, and the demographic history of the study population, and may therefore vary across populations and genomic regions. Nonetheless, current LAI methods achieve high accuracy in several well-characterized admixed populations, supporting the practical application of ancestry-aware analyses in these settings. Second, our analyses in AoU were restricted to HapMap3^15^ variants because local ancestry calls were available only at these markers currently; FELIX can be applied more broadly as denser calls become available. Future work should extend the framework to rare variants and gene-based association analysis, and to additional downstream applications of ancestry-resolved summary statistics.

As biobanks continue to expand, analyses based on discrete global ancestry groups can leave participants and ancestry-specific genetic information underused. By modeling ancestry at the haplotype level, FELIX enables full-cohort analysis that adapts to locus-specific genetic architecture, increasing power for locus discovery while producing more precise ancestry-resolved effect estimates that improve downstream polygenic prediction. Our findings establish local ancestry as a practical foundation for genetic analysis in increasingly admixed populations, allowing biobanks to expand diversity without sacrificing power, resolution, or scalability.

## Acknowledgements

We gratefully acknowledge *All of Us* participants for their contributions, without whom this research would not have been possible. We also thank the National Institutes of Health’s *All of Us* Research Program for making available the participant data examined in this study. This research has also been conducted using the UK Biobank Resource under Application Number 31063.

W.Z. is supported by the National Human Genome Research Institute of the National Institutes of Health under award number R00HG012222 and R01HG014518.

E.G.A. is supported by R01HG012869.

K.Y. is supported by the National Institute of Mental Health of the National Institutes of Health under award number K99MH135172.

B.L.G, B.M.N and K.J.K. are supported by the Novo Nordisk Foundation (NNF21SA0072102).

Y.W. is supported by NIH National Human Genome Research Institute K99 award under K99HG013969.

## Author information

These authors jointly supervised this work: Dr. Elizabeth Atkinson and Dr. Wei Zhou.

## Contributions

L.H., K.Y., E.G.A, and W.Z developed the FELIX framework and designed the computational analyses. K.Y. developed the FELIXla format. L.H. and T.T. designed the simulation pipeline. L.H. performed the benchmarking, association, and polygenic score real data analyses. Y. W. and W.Z. designed the polygenic score evaluation. B.L.G., W.L., and K.J.K. contributed to the comparison with the All-by-All project results. R.M., Z.W., and K.H. performed local ancestry inference in All of Us. A.R.M. and B.P. contributed to the local ancestry inference in All of Us. H.H., B.M.N., and M.J.D. contributed analytical and interpretation suggestions. W.Z. and E.G.A provided overall supervision for the project as co-corresponding authors. L.H., K.Y., and W.Z. wrote the original draft of the manuscript. All authors edited and reviewed the manuscript.

## Data Availability

This study used data from the All of Us Research Program’s Controlled Tier Dataset version 7 (GRCh38-aligned) available to authorized users on the Researcher Workbench. UK Biobank is an open-access resource. Applications to access the data from bona fide researchers can be made at https://www.ukbiobank.ac.uk/enable-your-research/apply-for-access. All relevant GWAS results (local-ancestry inferred summary statistics in this study) are available through Zenodo.

## Code Availability

FELIX is implemented as an open-source R method available at: https://github.com/ZhouLabGenetics/FELIX.

Code to reproduce analyses included in the manuscript can be found at: https://github.com/ZhouLabGenetics/FELIX_manuscript_code.

## ONLINE METHODS

### Ethics statement

This study analyzed de-identified participant data accessed through the All of Us Research Program Researcher Workbench and from the UK Biobank database. All participants provided informed consent for participation and use of their data for research. The UK Biobank received approval from the□North West Multi-centre Research Ethics Committee□as a□research tissue bank, allowing approved researchers to conduct analyses under this overarching ethics approval. Analyses regarding All of Us participants were performed in strict compliance with the All of Us Data User Code of Conduct and the Data and Statistics Dissemination Policy. No attempts were made to re-identify participants, and all reported findings contain aggregate data only. All analyses were performed in accordance with the relevant institutional and data-access requirements.

### Overview of the FELIX workflow

FELIX operates on phased^36^ genotypes and matched local ancestry calls. FELIX assumes default FLARE^8^ output to call ancestry-specific genotypes for each genetic marker in all samples. For individual *i*, variant *m*, haplotype *h* ∈ {1,2} and ancestry *k* ∈ {1… *K*.}, let *g*_*imh*_ ∈ {0,1} denote the alternative allele indicator and a_imh_∈ {1 · · · *K*} the inferred local ancestry. The ancestry-specific alternative allele dosage is 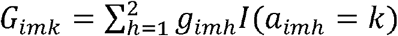, and the corresponding local ancestry haplotype count is 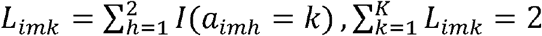. The aggregate alternative allele dosage is 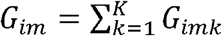. FELIXla stores *G*_*imk*_ and *L*_*imk*_ in a compact, variant-queryable representation. FELIXassoc uses these quantities for shared-effect and ancestry-specific association testing.

### FELIXla: a haplotype-resolved, local-ancestry-aware genotype storage format

Ancestry-specific dosages can be reconstructed from phased genotypes and local ancestry files but recomputing them for every variant and phenotype can be expensive at biobank scale. FELIXla is a binary format that stores phased alleles and their inferred local ancestries together and returns ancestry-specific dosages by direct query. Each haplotype is indexed explicitly, the alternate-allele state of each haplotype is stored as one bit, and local ancestry is stored as ancestry-specific haplotype masks under the same index. The ancestry K dosage at a variant is then a bitwise intersection of the genotype vector and the ancestry K mask. Because local ancestry is piecewise constant along a chromosome, FELIXla stores ancestry transition points rather than a label at every variant, and it uses dense bit vectors for common variants and sparse carrier lists for rare variants. The bit-level layout, the transition encoding and the chunked input and output are described in **Supplementary Note**.

### FELIXassoc statistical model for association testing

FELIXassoc extends the SAIGE^18^ generalized linear mixed-model framework to ancestry-resolved genotypes. For individual *i*, let Y_i_ denote the phenotype and C_i_ a vector of fixed-effect covariates that includes age, sex, genetic principal components and an intercept. The null model is 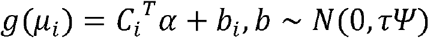, where *g*(·) is the identity link for quantitative traits and the logit link for binary traits, *µ*_*i*_ is the mean of phenotype under the null hypothesis, *Ψ* is an ancestry-adjusted sparse genetic relationship matrix and *τ* is the polygenic genetic variance parameter. The sparse GRM accounts for cryptic relatedness while allowing participants to span a continuous global-ancestry distribution. In this analysis, we constructed with FastSparseGRM^17^, which projects out ancestry before thresholding, so that relatedness rather than shared ancestry is retained. Fixed effects and variance components are estimated once per phenotype under the null hypothesis of no association using the penalized quasi-likelihood^37^ and average-information restricted maximum-likelihood algorithm^38^, inheriting from SAIGE^18^ framework, to iteratively estimate 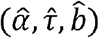. The fitted null model is reused for all variants and both association tests.

#### Homogeneous and heterogeneous association tests

Under the homogeneous (shared-effect) model, the aggregate dosage G_im_ has one marginal effect across ancestral backgrounds, 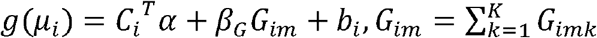, and FELIXassoc tests *H*_0_: *β*_*G*_ = 0 with a one-degree-of-freedom score test. With the summed dosage vector g, the one-dimentional score is 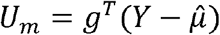. This test has the same structure as the standard SAIGE single-variant test but uses the full cohort without assigning participants to discrete ancestry groups. The haplotype effects across ancestries can be accounted for by ancestry PCs, which are usually included as covariates in *C*_*i*_.

Under the heterogeneous (ancestry-specific) model, each background carries its own per-allele effect, and local ancestry enters as a covariate, 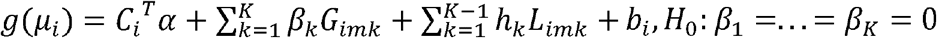, where *β*_*k*_ is the per-allele effect on the ancestry k background, and h_k_ is the effect of the ancestry k haplotype count. The haplotype term adjusts for admixture linkage disequilibrium and is included adaptively (see *Conditioning on local ancestry haplotype counts* section below). In a study of sample size N, let G_m_ be the *N* × *K*. matrix whose columns hold the ancestry-specific dosages G_imk_ at variant m, and 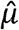 the fitted null mean. The K-vector score 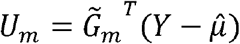 is the difference between observed and null-expected ancestry-specific allele counts, where 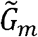 is G_m_ with the covariates projected out. Its exact variance under *H*_0_ is 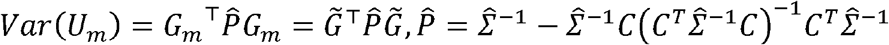, where p is the null-model projections, *Σ*. is the marginal covariance of the working vector. The heterogeneous test statistic is *T*_*het,m*_ = *U*_*m*_^*T*^*Var(U*_*m*_)^-1^*U*_*m*_ is compared with a chi-squared distribution on degrees of freedom equal to the rank of *V*_*m*_. Computing *V*_*m*_ exactly for every variant would require expensive operations on the full *N* × *N* covariance. FELIXassoc instead uses the variance-ratio approximation of SAIGE^18^, estimating a ratio from a subset of variants and applying it genome-wide so that score statistics are formed from covariate-projected dosages without repeated inversion. The variance-ratio derivation is given in **Supplementary Note**.

#### Saddlepoint approximation for imbalanced binary traits

For binary traits with highly imbalanced case-control ratios, the normal approximation to the score-statistic distribution can inflate type I error, particularly for low-frequency variants (**Supplementary Figure 3**). FELIXassoc therefore applies the Saddlepoint approximation^19^ (SPA). For the shared-effect test, SPA is applied to the one-dimensional score statistic. For the ancestry-specific test, SPA is applied to each marginal ancestry-specific score statistic, and the ratio of SPA-adjusted to normal-approximation variance rescales the corresponding diagonal and covariance terms of *V*_*m*_ before the multi-degree-of-freedom statistic is formed. As in SAIGE^18^, SPA is applied only when the absolute normal-approximation z-score exceeds 2, where it is numerically stable and beneficial. The variance-rescaling expressions are given in **Supplementary Note**.

#### Adaptive combination of the homogeneous and heterogeneous tests

Let P_hom_ and P_het_ denote the shared-effect and ancestry-specific p values. FELIXassoc combines them with the Cauchy combination test^14^, *T*_*CCT*_ = 0.5 × [*tan* {0.5 – *p*_*hom*_) *π*} + *tan*{0.5 - *p*_*het*_)*π*}], *P*_*CCT*_ = 0.5 – *arctan*(*T*_*CCT*_)/ *π*. Equal weights are assigned to the two tests. The Cauchy combination test^14^ is valid under arbitrary dependence between the component statistics and needs no estimate of their correlation. The combined test therefore retains the efficiency of the shared-effect model where effects are concordant and preserves sensitivity where effects are ancestry-dependent, with power tracking the better-fitting model at each locus (**Supplementary Figure 4**).

#### Conditioning on local ancestry haplotype counts

At certain loci, the local ancestry haplotype count is strongly associated with both dosage and phenotype, for example at *HBB* region when analyzing sickle-cell disease. Marginal ancestry-specific tests can then return mirrored effect estimates, because the dosage assigned to one ancestry is negatively correlated with the dosage or local ancestry assigned to another. FELIXassoc therefore applies an adaptive conditioning procedure. At each variant, the association between the phenotype and each local ancestry haplotype count L_imk_ is first evaluated with a marginal score test, with SPA applied for imbalanced binary traits. If a haplotype-count association reaches user-specified haplotype significance threshold (default *p* <5 × 10^−6^), the corresponding counts are included in a conditional score test that separates the allelic association from the association attributable to local ancestry exposure. Where local ancestry carries little information about dosage, the marginal test is reported, so that ancestry-specific variants that are too rare for ancestry to predict carrier status are not conditioned unnecessarily. The joint covariance used for conditioning is given in **Supplementary Note**.

### Simulation of three-way admixed cohorts for FELIXassoc

We simulated a three-way African-European-Indigenous American (AFR-EUR-AMR) admixed cohort to evaluate calibration and power under multi-way admixture. The cohort comprised N = 10,000 individuals: 5,000 unrelated and 5,000 related (500 three-generation families of 10 retained descendants each, pedigree diagram as **Supplementary Figure 10**). Genotypes for related individuals were produced by gene-dropping to preserve family relationships. Six unrelated founders each drew an independent Dirichlet distribution. Ancestry-specific dosages were computed as *DS*_*k*_ = *I*(*la*_l_ = *k*) × *hap*_l_ + *I*(*la*_2_ = *k*) × *hap*_2_, and local ancestry haplotype counts as *ANC*_*k*_ = *I*(*la*_l_ = *k*) + *I*(*la*_2_ = *k*). Ancestry-specific allele frequencies were generated using a three-population Balding-Nichols model, with per-population differentiation parameters *F*_*ST,AFR*_= 0.15, *F*_*ST,EUR*_ = 0.10, and *F*_*ST,AMR*_ = 0.15. These values, drawn from empirical estimates in continental populations^37–39^, yield induced pairwise *F*_*ST*_ values of approximately 0.15 (AFR-EUR), 0.19 (AFR-AMR), and 0.11 (EUR-AMR), consistent with published estimates. Individual-level global ancestry proportions were drawn from a Dirichlet(1.0, 5.5, 3.5) distribution, producing a Latino-like admixture profile with mean proportions of approximately 10% AFR, 55% EUR, and 35% AMR.

#### Continuous phenotypes were generated as

*γ*_*i*_ = *γ*_*AFR*_ × *Admprop*_{*i AFR*}_+ *γ*_*AMR*_ × *Admprop*_*{i,AMR}*_ + *b*_*i*_ + *ϵ*_*i*_ ∑(*β*_*causal,k*_ ×*G*_*i,c*_^(a)^), where *Admprop*_*{i,a}*_ denotes participant *i*’s genome-wide proportion of ancestry *a* (European is the reference to avoid collinearity), *γ*_*AFR*_ and *γ*_*AMR*_ are the corresponding mean effects, G_*i,c*_^(*a*)^ is the allele dosage at causal variant *r* carried on the ancestry *a* background, *β*_*causal,k*_ is the causal effect on that background, *b* ∼ *MVN*(0,*σ*_*g*_^2^ × Ψ) is the polygenic random effect with Ψ = 2 × kinship and,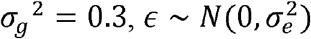 with, *σ*_*e*_^2^ = 0.7. Binary phenotypes were generated from the corresponding logistic mixed model, with intercept *β*_0_ calibrated via root-finding to achieve target prevalences of 1% and 10%.

For type I error assessment, *β*_*causal,k*_ = 0 for all k, and 10 null phenotype replicates were generated per trait, each tested against approximately 1 million variants across 20 simulated chromosomes. For power assessment, a single causal variant was selected from the first simulated chromosome with MAF closest to 0.20 (common) or 0.03 (low-frequency) and nonzero variance in all three ancestry-specific dosages. Two genetic architectures were evaluated: (i) shared effects *β*_*causal,AFR*_ = *β*_*causal,EUR*_ *β*_*causal,AMR*_ = *B*; and (ii) asymmetric heterogeneous effects *β*_*causal,AFR*_ = 0, *β*_*causal,EUR*_ 0.5B, *β*_*causal,AMR*_ = *B*. Simulation was performed with 100 replicate seeds per condition. Power was computed as the fraction of replicates in which the causal variant’s p-value fell below the significance threshold (**Supplementary Figure 4**).

### Local ancestry inference on HapMap3 variants in All-of-Us

Further details on local ancestry inference in AoU v7 have been previously reported^39^. Briefly, local ancestry inference in AoU was performed using RFMix^41^ v2 with default parameters (eight generations since admixture) with reference super-populations from 1000 Genomes Project Phase 3^40^ (African: AFR, European: EUR, East Asian: EAS, South Asian: SAS) and samples from Lima, Peru (Indigenous American). Local ancestry was inferred at HapMap3^15^ SNPs, yielding per-haplotype ancestry calls at 1,042,379 variants. The inferred per-haplotype ancestry calls, together with the phased genotypes, provided the ancestry-specific dosages and local ancestry haplotype counts stored in the FELIXla format.

### Phenotype, meta-analysis comparison and precision

We analyzed 24 quantitative and binary phenotypes covering biomarkers, lab and physical measurements, and disease outcomes (as defined using phecode^41^ and phecodeX^42^ curation; **Supplementary Table 3**). We compared FELIXassoc with the fixed-effect All-by-All global-ancestry meta-analysis on the same AoU^4^ release, restricted to the shared variants. Signals whose lead variants were within +/-500 kb were treated as the same region. Near-miss signals were defined as loci with p-value between 5 × 10^-8^ to 5 × 10^−6^ in the comparison.

### Empirical polygenic score prediction

To assess the downstream value of full-cohort summary statistics, we constructed PRS from AoU^2,4^ summary statistics and evaluated prediction in an independent UK Biobank^16^ cohort. For each of the 11 quantitative traits examined in FELIXassoc (nine biomarkers, BMI and height), we constructed three PRSs based on three sets of discovery summary statistics: FELIXassoc full cohort estimate analyzed jointly for all local ancestries, All-by-All^4^ global-ancestry-based meta-analysis, and All-by-All global-ancestry-stratified estimate. Posterior effect sizes were estimated through PRS-CS^33^ using the European Linkage-disequilibrium reference panel and the default, fully Bayesian global shrinkage parameter. Scores were computed with PLINK2^43^. Prediction was evaluated in the four Pan-UK Biobank^34^ validation cohorts (EUR/AFR/EAS/CSA, per-trait sample size in **Supplementary Table 9**) to determine R^2^ from a regression of the rank-inverse-normalized phenotype on the score. Accuracy was quantified as the incremental coefficient of determination (incremental R^2^). For each score we fitted a covariate-only model including 20 genetic principal components, age, sex, age×sex, age^2^ and age^2^×sex, and a full model adding the polygenic score, and took the difference in R^2^ between them to report the phenotypic variance explained by the score beyond the covariates. 95% confidence intervals for each incremental R^2^ and for the local-global difference were obtained by a nonparametric bootstrap that resampled the validation individuals with replacement over 2,000 replicates, recomputing the statistic in each replicate and taking the 2.5th and 97.5th percentiles. The same resampled individuals were used for both scores within each replicate, so the difference is a paired bootstrap and its interval accounts for the correlation between the two scores.

---

**Supplementary Figure 1.**
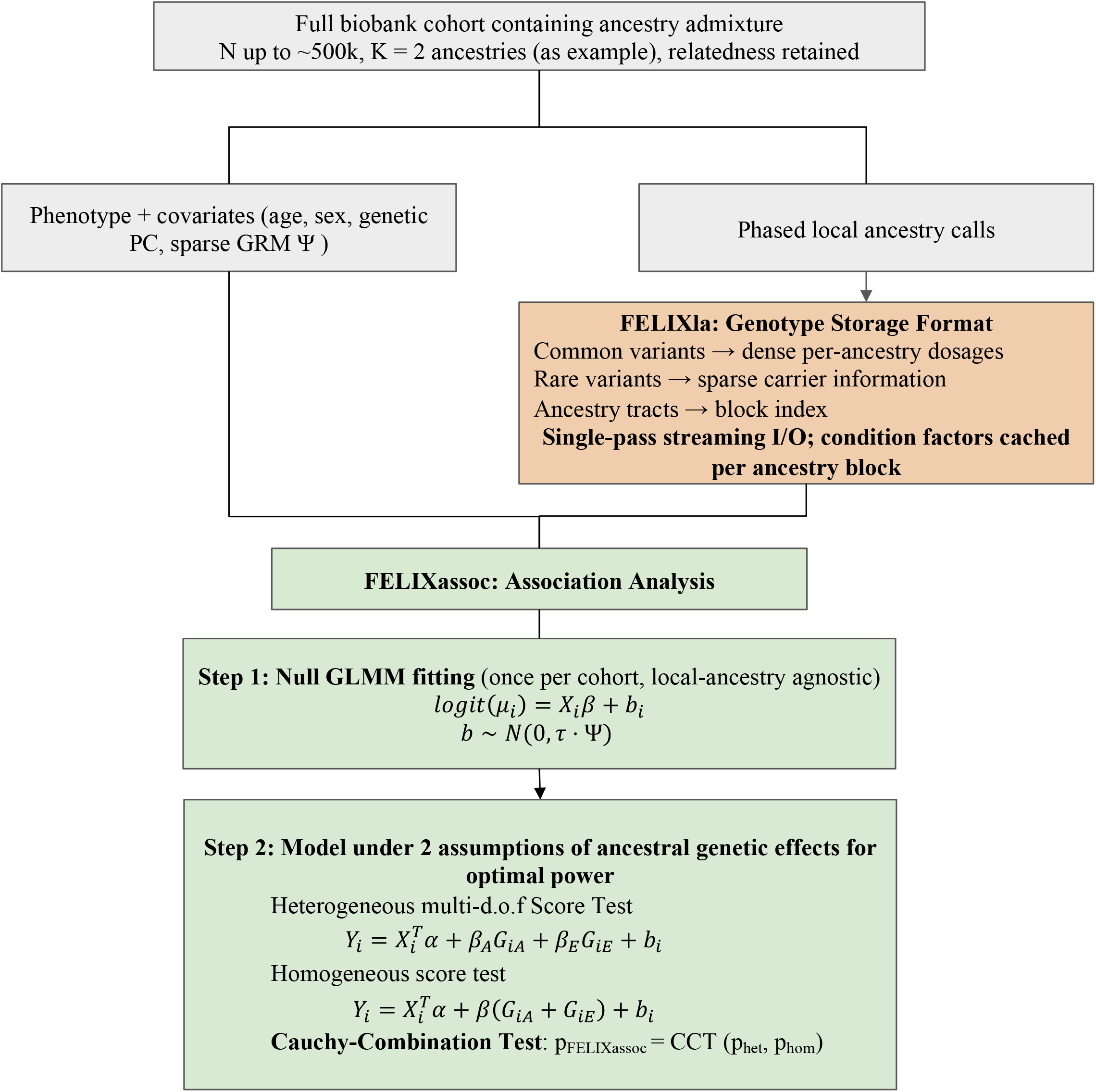
FELIX method schematic and storage format. Key steps, input arguments and parameters estimated of the FELIX workflow. Details can be found in the **Online Methods** and **Supplementary Note**. Denotation: Y, phenotype vector; G, aggregate (summed-across-ancestries) alternative-allele dosage used by the homogeneous test;*G*_*iA*_, ancestry-A alternative-allele dosage; μ, fitted mean from the Step 1 null model; Ψ, sparse genetic relationship matrix; τ, polygenic variance parameter; Ψ, number of local ancestries; ß, per-allele effect; d.o.f., degrees of freedom; CCT, Cauchy combination test.

**Supplementary Figure 2.**
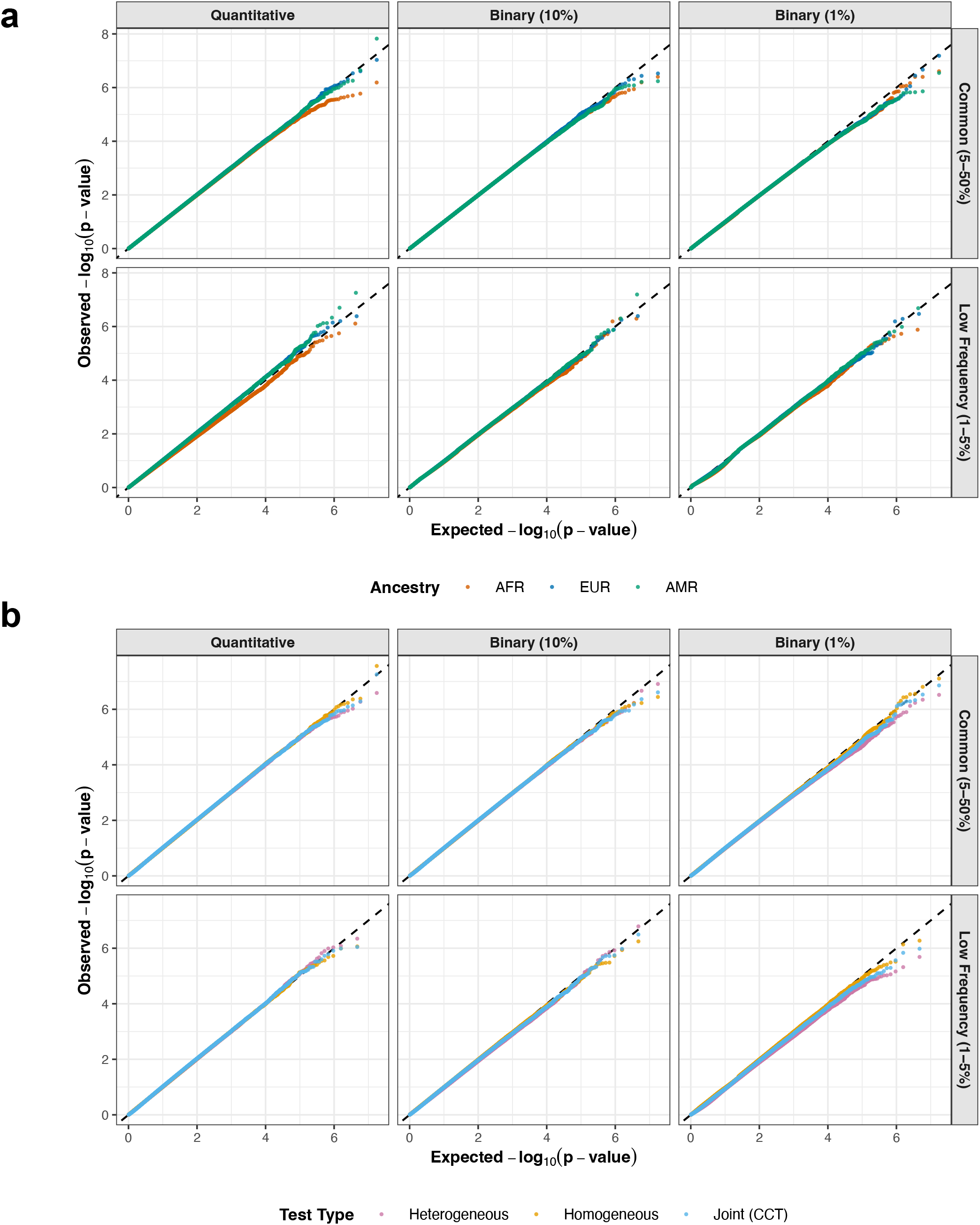
Type I error under multi-way simulated admixture. Empirical type I error for each local-ancestry component and for the homogeneous, heterogeneous, and Cauchy-combined tests, from 10^7^ null simulations in a three-way admixed cohort (*N* = 10,000). Test evaluated on quantitative traits and on binary traits at 10% and 1% prevalences, and across frequency bins. **a**, ancestry-specific calibration; **b**, test-specific calibration. Calibration is maintained across variant-frequency bins, trait types, and degrees of case-control imbalance.

**Supplementary Figure 3.**
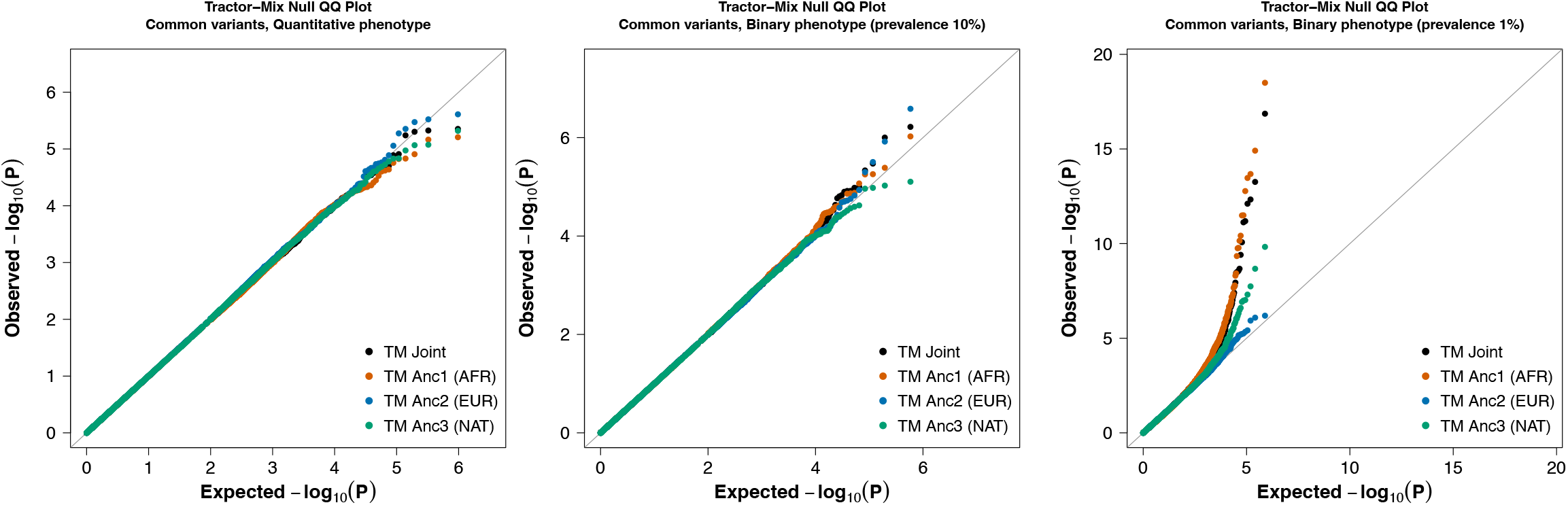
Type I error of Tractor-Mix under the multi-way simulated admixture. Under the same simulation settings, Tractor-Mix is calibrated for common variants in quantitative traits and 10%-prevalence binary traits. But Tractor-Mix shows inflated type I error for 1% prevalence binary traits, in both its joint and per-ancestry-tract statistics.

**Supplementary Figure 4.**
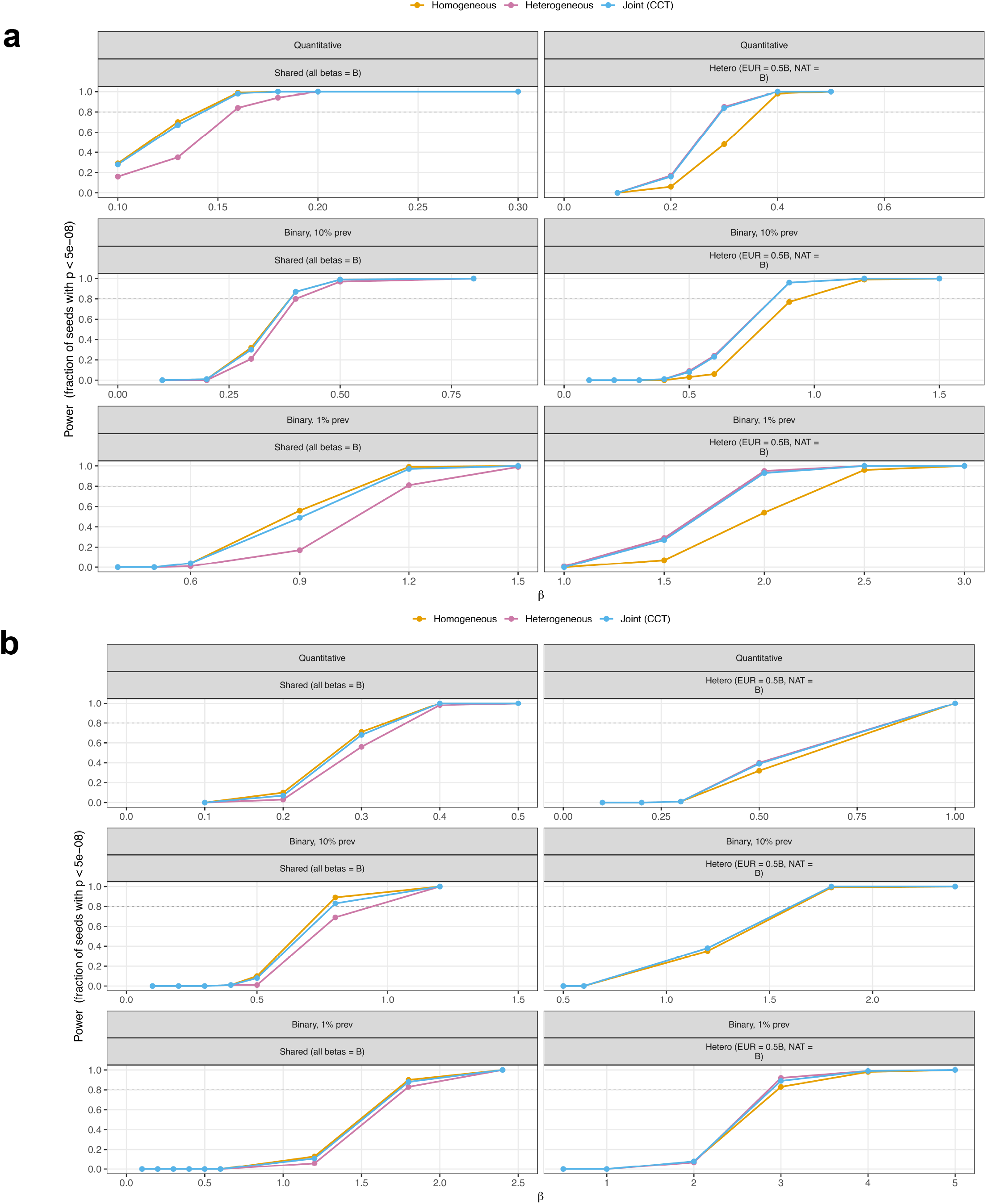
Power tracks the locus-optimal model under multi-way simulated admixture. The power of all component tests in FELIXassoc is evaluated under homogeneous (shared-effect) and asymmetric heterogeneous architectures. The homogeneous test is most powerful under shared effects, the heterogeneous test under divergent effects, and the Cauchy-combined test matches the locus-optimal test across architectures. **a**, Common variants (MAF > 5%). **b**, Lower-frequency variants (1% < MAF < 5%). MAF, minor allele frequency.

**Supplementary Figure 5.**
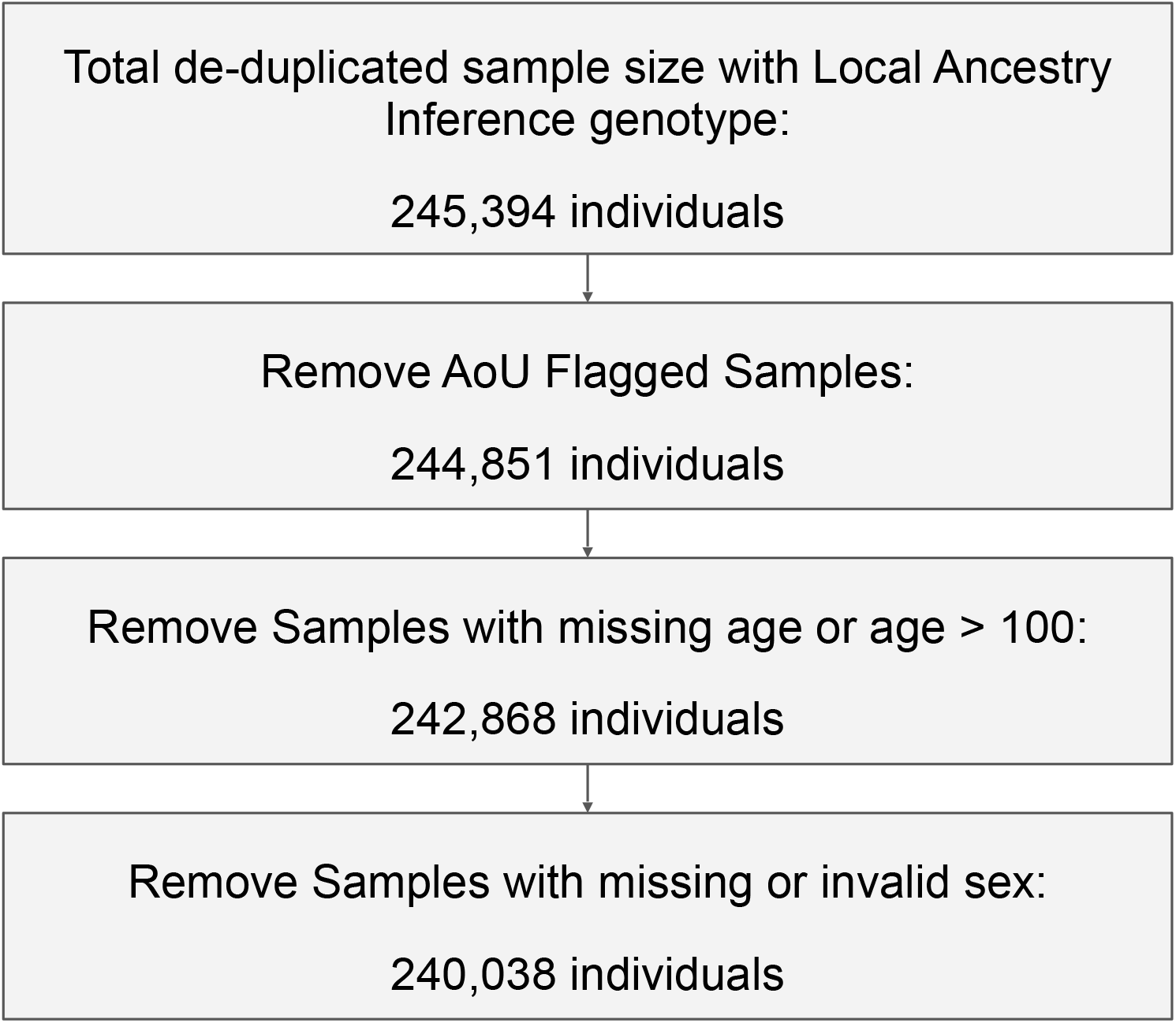
Sample quality control workflow in All of Us. Sample filtering steps applied to the All of Us cohort before FELIX analysis, with the number of participants remaining after each step.

**Supplementary Figure 6.**
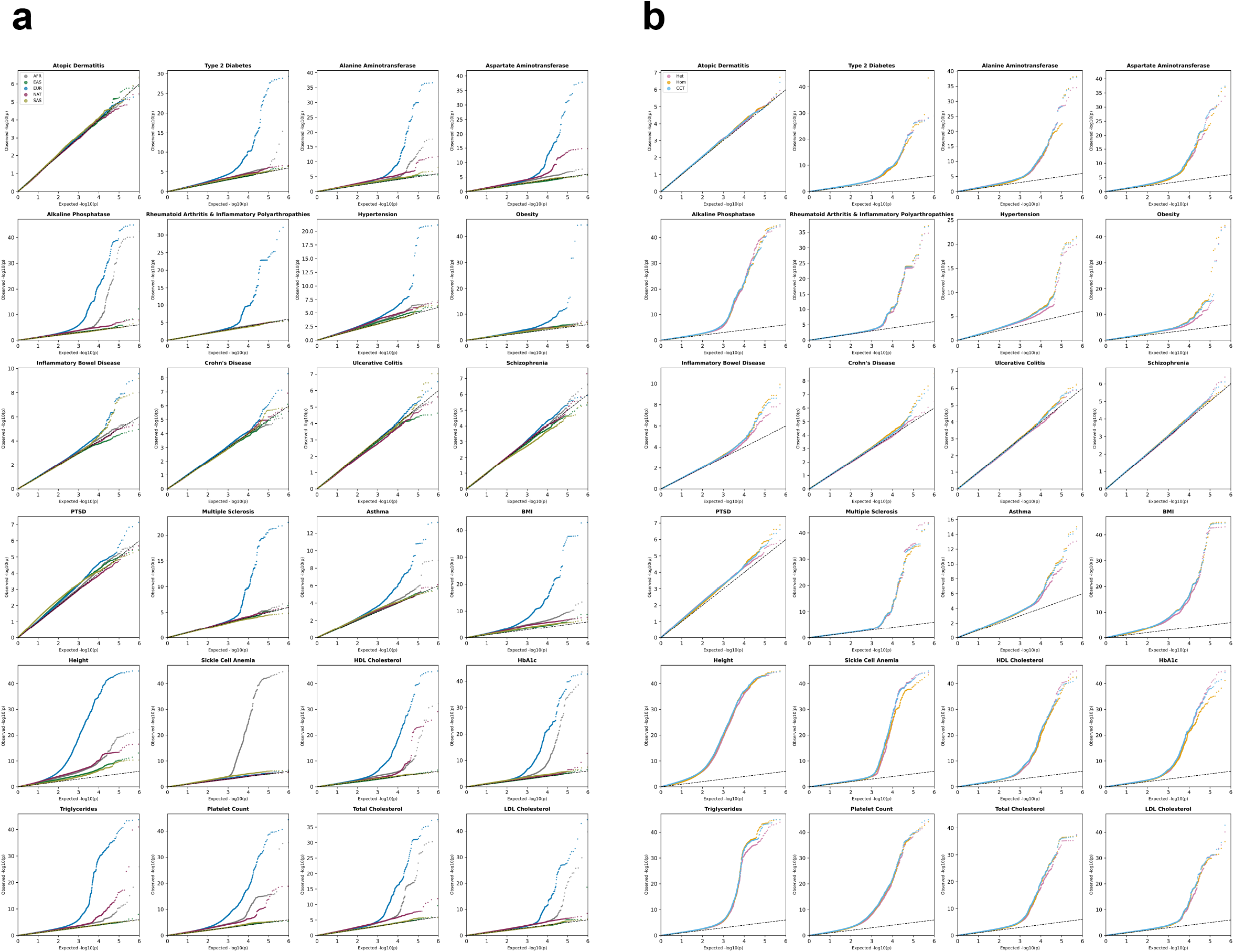
Empirical calibration in All of Us across phenotypes. Quantile-quantile plots of FELIXassoc in the five-way admixed All of Us cohort analysis show no systematic inflation. **a**, By-local-ancestry component FELIXassoc summary statistics. **b**, By-test component FELIXassoc summary statistics (homogeneous, heterogeneous, Cauchy-combined).

**Supplementary Figure 7.**
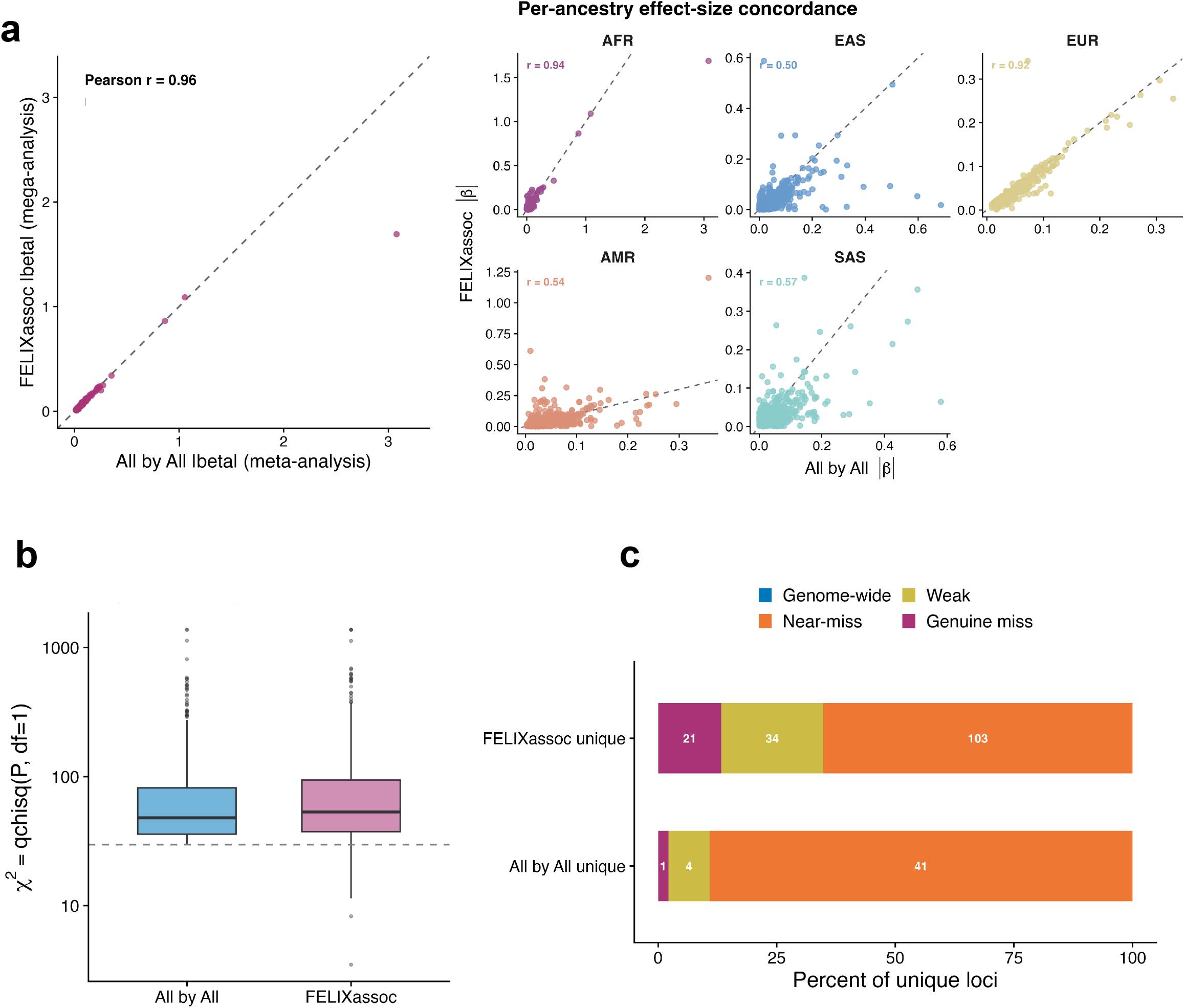
High concordance of FELIXassoc with global-ancestry meta-analysis results (All-by-All). **a**, Highly concordant effect sizes comparing FELIXassoc homogeneous test results and All-by-All meta-analysis summary statistics across All-by-All meta-analysis significant loci, and the ancestry-matched effect sizes concordance shown in the right panel. **b**, At the genome-wide significant baseline loci, FELIXassoc displays higher chi-squared statistics compared to global-ancestry-based meta-analysis. **c**, Loci reaching genome-wide significance in only FELIXassoc or only All-by-All.

**Supplementary Figure 8.**
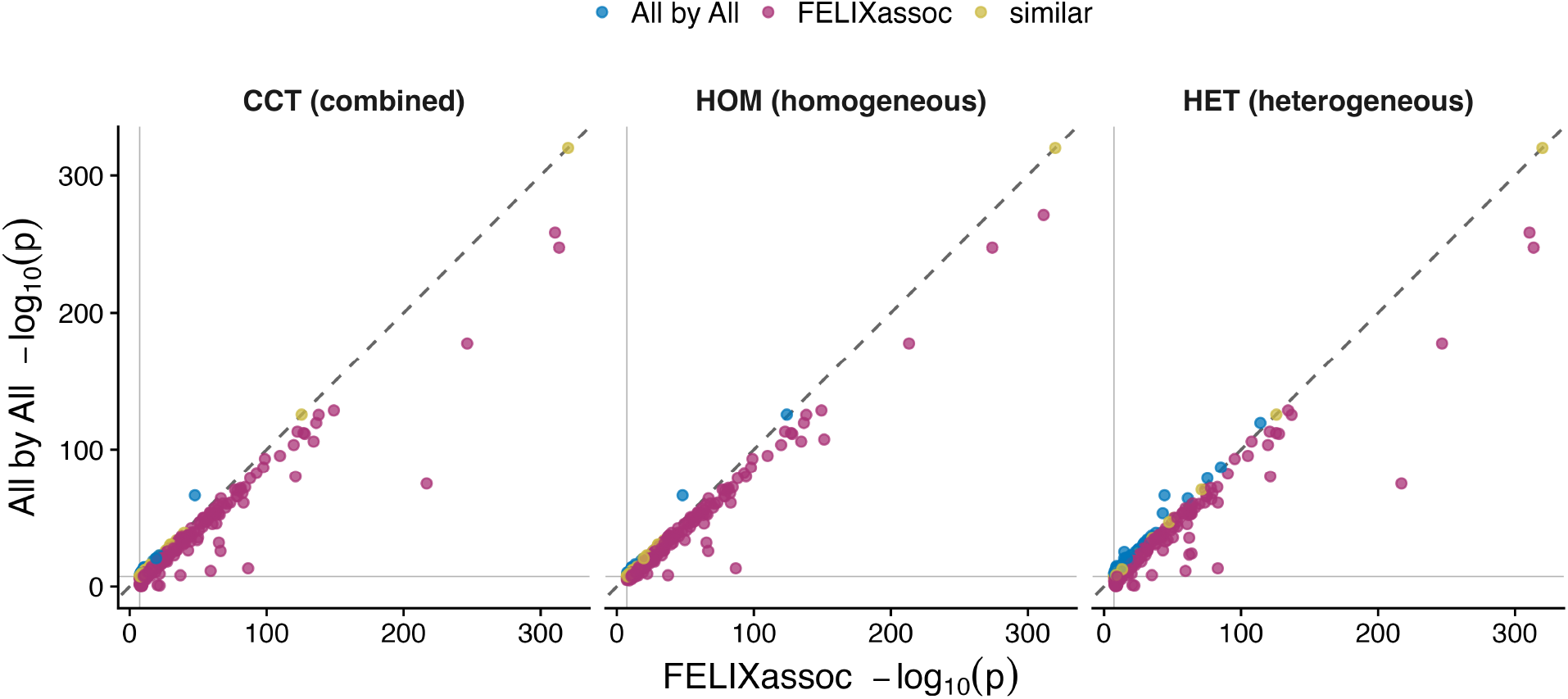
Stronger signal at shared significant loci. Signal strength at loci significant in both FELIXassoc and All-by-All analyses. Overall, the local-ancestry-based FELIXassoc tests show a stronger association than the global-ancestry meta-analysis (blue: All-by-All produced a stronger signal at the shared significant locus; purple: FELIXassoc produced a stronger signal at the shared significant locus; yellow: All-by-All and FELIXassoc produced similarly strong signals at the shared significant locus).

**Supplementary Figure 9.**
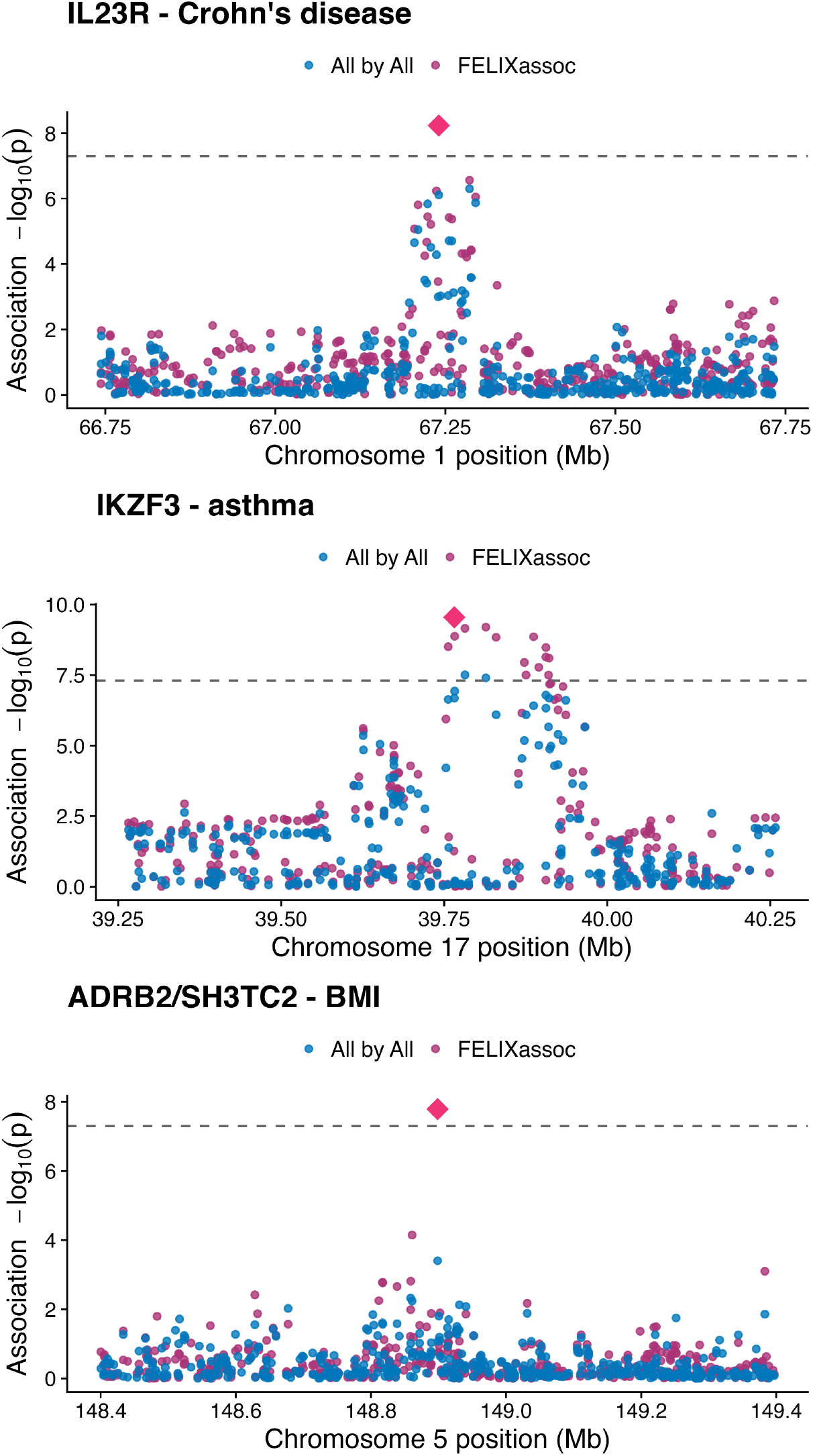
Locus-level view of the haplotype-recovery discovery mechanism. LocusZoom plots contrasting FELIXassoc with global-ancestry meta-analysis results, illustrating how recovering under-represented ancestry-specific haplotypes increases statistical power for discovery.

**Supplementary Figure 10.**
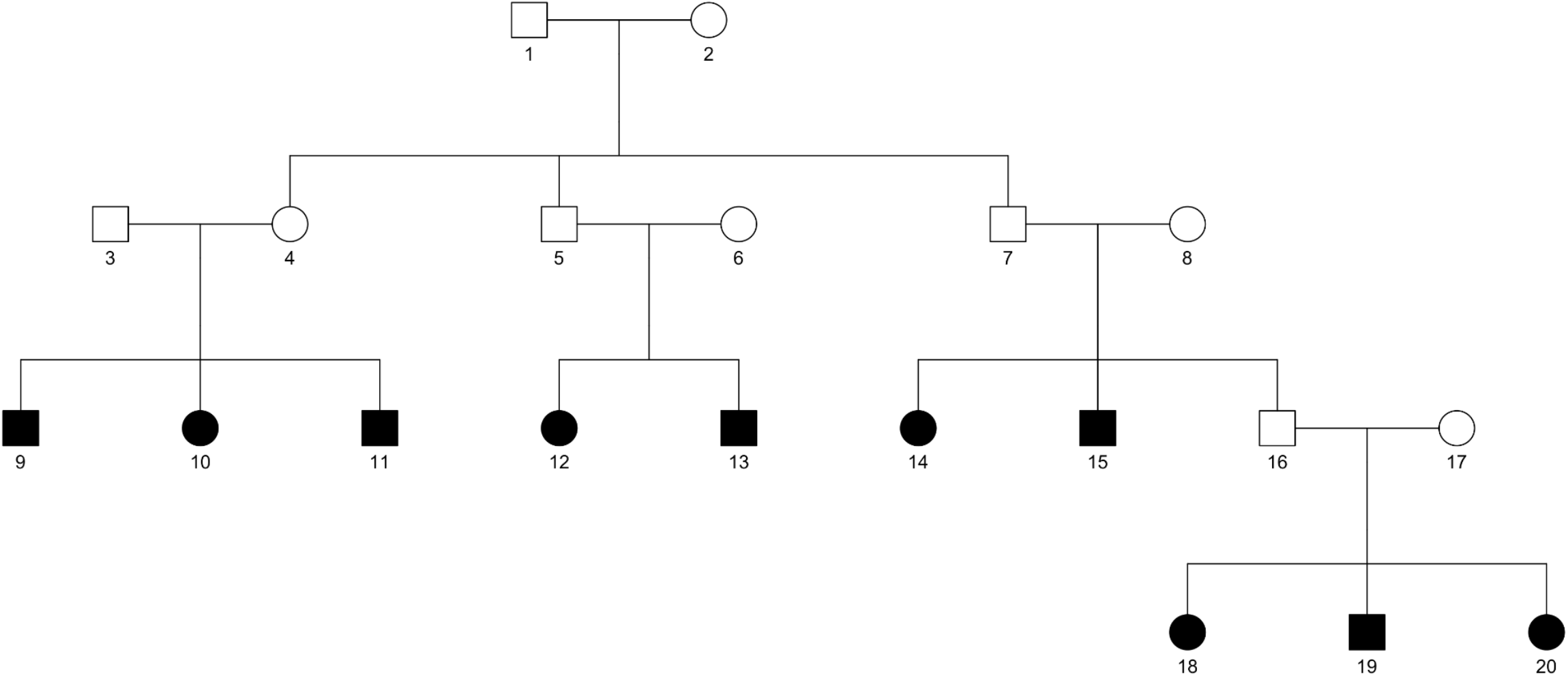
Pedigree structure of the three-generation families in simulations. Squares denote males, circles denote females; filled symbols indicate the 10 descendants retained per family for downstream analyses.

## References

1. Zhou, W. et al. Global Biobank Meta-analysis Initiative: Powering genetic discovery across human disease. Cell Genom. 2, 100192 (2022).

2. All of Us Research Program Investigators et al. The “All of Us” Research Program. N. Engl. J. Med. 381, 668–676 (2019).

3. Gaziano, J. M. et al. Million Veteran Program: A mega-biobank to study genetic influences on health and disease. J. Clin. Epidemiol. 70, 214–223 (2016).

4. Lu, W. et al. Systematic common and rare variant association testing in 392,030 whole genomes in All of Us. medRxiv (2026) doi:10.64898/2026.05.08.26350964.

5. Wojcik, G. L. et al. Genetic analyses of diverse populations improves discovery for complex traits. Nature 570, 514–518 (2019).

6. Curtis, D. Polygenic risk score for schizophrenia is more strongly associated with ancestry than with schizophrenia. Psychiatr. Genet. 28, 85–89 (2018).

7. Martin, A. R. et al. Clinical use of current polygenic risk scores may exacerbate health disparities. Nat. Genet. 51, 584–591 (2019).

8. Browning, S. R., Waples, R. K. & Browning, B. L. Fast, accurate local ancestry inference with FLARE. Am. J. Hum. Genet. 110, 326–335 (2023).

9. Atkinson, E. G. et al. Tractor uses local ancestry to enable the inclusion of admixed individuals in GWAS and to boost power. Nat. Genet. 53, 195–204 (2021).

10. Tan, T. et al. Extending Genome-Wide Association Studies to admixed cohorts with high degrees of relatedness. medRxiv (2025) doi:10.1101/2025.05.27.25328444.

11. Hou, K., Bhattacharya, A., Mester, R., Burch, K. S. & Pasaniuc, B. On powerful GWAS in admixed populations. Nat. Genet. 53, 1631–1633 (2021).

12. Hou, K. et al. Causal effects on complex traits are similar for common variants across segments of different continental ancestries within admixed individuals. Nat. Genet. 55, 549–558 (2023).

13. Mester, R. et al. Impact of cross-ancestry genetic architecture on GWASs in admixed populations. Am. J. Hum. Genet. 110, 927–939 (2023).

14. Liu, Y. & Xie, J. Cauchy combination test: a powerful test with analytic p-value calculation under arbitrary dependency structures. J. Am. Stat. Assoc. 115, 393–402 (2020).

15. International HapMap 3 Consortium et al. Integrating common and rare genetic variation in diverse human populations. Nature 467, 52–58 (2010).

16. Sudlow, C. et al. UK biobank: an open access resource for identifying the causes of a wide range of complex diseases of middle and old age. PLoS Med. 12, e1001779 (2015).

17. Lin, X., Dey, R., Li, X. & Li, Z. Scalable analysis of large multi-ancestry biobanks by leveraging sparse ancestry-adjusted sample-relatedness. Res. Sq. (2024) doi:10.21203/rs.3.rs-5343361/v1.

18. Zhou, W. et al. Efficiently controlling for case-control imbalance and sample relatedness in large-scale genetic association studies. Nat. Genet. 50, 1335–1341 (2018).

19. Daniels, H. E. Saddlepoint Approximations in Statistics. Ann. Math. Stat. 25, 631–650 (1954).

20. Balding, D. J. & Nichols, R. A. A method for quantifying differentiation between populations at multi-allelic loci and its implications for investigating identity and paternity. Genetica 133, 107–107 (2008).

21. Svishcheva, G. R., Axenovich, T. I., Belonogova, N. M., van Duijn, C. M. & Aulchenko, Y. S. Rapid variance components-based method for whole-genome association analysis. Nat. Genet. 44, 1166–1170 (2012).

22. Loh, P.-R. et al. Efficient Bayesian mixed-model analysis increases association power in large cohorts. Nat. Genet. 47, 284–290 (2015).

23. Jiang, L. et al. A resource-efficient tool for mixed model association analysis of large-scale data. Nat. Genet. 51, 1749–1755 (2019).

24. Bryc, K., Durand, E. Y., Macpherson, J. M., Reich, D. & Mountain, J. L. The genetic ancestry of African Americans, Latinos, and European Americans across the United States. Am. J. Hum. Genet. 96, 37–53 (2015).

25. Abdollahi, E., Tavasolian, F., Momtazi-Borojeni, A. A., Samadi, M. & Rafatpanah, H. Protective role of R381Q (rs11209026) polymorphism in IL-23R gene in immune-mediated diseases: A comprehensive review. J. Immunotoxicol. 13, 286–300 (2016).

26. Duerr, R. H. et al. A genome-wide association study identifies IL23R as an inflammatory bowel disease gene. Science 314, 1461–1463 (2006).

27. Romeo, S. et al. Genetic variation in PNPLA3 confers susceptibility to nonalcoholic fatty liver disease. Nat. Genet. 40, 1461–1465 (2008).

28. Wagenknecht, L. E. et al. Association of PNPLA3 with non-alcoholic fatty liver disease in a minority cohort: the Insulin Resistance Atherosclerosis Family Study. Liver Int. 31, 412–416 (2011).

29. Chen, L.-Z. et al. PNPLA3 I148M variant in nonalcoholic fatty liver disease: demographic and ethnic characteristics and the role of the variant in nonalcoholic fatty liver fibrosis. World J. Gastroenterol. 21, 794–802 (2015).

30. Kore, P. et al. Improved allele frequencies in gnomAD through local ancestry inference. bioRxivorg (2025) doi:10.1101/2024.10.30.620961.

31. Hardwick, R. J. et al. Haptoglobin (HP) and Haptoglobin-related protein (HPR) copy number variation, natural selection, and trypanosomiasis. Hum. Genet. 133, 69–83 (2014).

32. Kozlitina, J. et al. Plasma levels of risk-variant APOL1 do not associate with renal disease in a population-based cohort. J. Am. Soc. Nephrol. 27, 3204–3219 (2016).

33. Ge, T., Chen, C.-Y., Ni, Y., Feng, Y.-C. A. & Smoller, J. W. Polygenic prediction via Bayesian regression and continuous shrinkage priors. Nat. Commun. 10, 1776 (2019).

34. Karczewski, K. J. et al. Pan-UK Biobank genome-wide association analyses enhance discovery and resolution of ancestry-enriched effects. Nat. Genet. 57, 2408–2417 (2025).

35. US Census Bureau. 2020 Census illuminates racial and ethnic composition of the country. Census.gov https://www.census.gov/library/stories/2021/08/improved-race-ethnicity-measures-reveal-united-states-population-much-more-multiracial.html (2021).

36. Delaneau, O., Marchini, J. & Zagury, J.-F. A linear complexity phasing method for thousands of genomes. Nat. Methods 9, 179–181 (2011).

37. Golan, D., Lander, E. S. & Rosset, S. Measuring missing heritability: inferring the contribution of common variants. Proc. Natl. Acad. Sci. U. S. A. 111, E5272–81 (2014).

38. Gilmour, A. R., Thompson, R. & Cullis, B. R. Average information REML: An efficient algorithm for variance parameter estimation in linear mixed models. Biometrics 51, 1440 (1995).

39. Mandla, R. et al. Large-scale admixture mapping in the All of Us Research Program improves the characterization of cross-population phenotypic differences. medRxiv (2025) doi:10.1101/2025.04.02.25325115.

40. 1000 Genomes Project Consortium et al. A global reference for human genetic variation. Nature 526, 68–74 (2015).

41. Wei, W.-Q. et al. Evaluating phecodes, clinical classification software, and ICD-9-CM codes for phenome-wide association studies in the electronic health record. PLoS One 12, e0175508 (2017).

42. Shuey, M. M. et al. Next-generation phenotyping: introducing phecodeX for enhanced discovery research in medical phenomics. Bioinformatics 39, btad655 (2023).

43. Chang, C. C. et al. Second-generation PLINK: rising to the challenge of larger and richer datasets. Gigascience 4, 7 (2015).

44. NIAID Visual & Medical Arts. Macrophage. nih.gov (2024).

