## Supplementary Notes for "A unified framework for local-ancestry-aware genetic association analysis across biobanks"

### Supplementary Note

#### Supplementary Note 1. FELIXla storage format

##### 1.1 Haplotype indexing and bitwise dosage query

FELIX operates on ancestry-specific dosages, which can be reconstructed from phased genotypes[1] and local ancestry calls[2,3] but are expensive to store distinctly for every variant at biobank scale. FELIXla is a binary format that stores phased alleles and their inferred local ancestries together and returns ancestry-specific dosages by direct query. Each of the $2N$ haplotypes of $N$ diploid individuals is indexed explicitly. The alternative allele state of each haplotype is stored as a single bit $g_{imh} \in \{0, 1\}$. Local ancestry is stored as ancestry-specific haplotype masks under the same index: the ancestry-$a$ mask bit for haplotype $h$ of individual $i$ at variant $m$ is $\mathbb{1}(\text{haplotype } h \text{ is ancestry } a)$. The ancestry-$a$ alternative allele dosage and haplotype count at a variant are then obtained by a bitwise intersection of the genotype bits with the ancestry-$a$ mask,

$$G_{im}^{(a)} = \sum_{h=1}^{2} g_{imh}\, \mathbb{1}(\text{haplotype } h \text{ is ancestry } a), \qquad L_{im}^{(a)} = \sum_{h=1}^{2} \mathbb{1}(\text{haplotype } h \text{ is ancestry } a), \qquad \text{(S1)}$$

with $\sum_a L_{im}^{(a)} = 2$ and aggregate dosage $G_{im} = \sum_a G_{im}^{(a)}$. Across a cohort, the intersection is a population-scale bitwise and followed by a population count, so ancestry-resolved dosages are recovered without re-parsing phased VCF files.

##### 1.2 Transition encoding and hybrid dense/sparse storage

Because local ancestry is piecewise constant along a chromosome, FELIXla stores ancestry transition points per haplotype rather than repeating an ancestry label at every variant, so the ancestry mask for any variant is recovered from the enclosing ancestry block. The format also adapts to the allele-frequency spectrum. Common variants are stored as dense bit vectors across haplotypes, whereas rare variants are stored as sparse lists of the haplotypes that carry the alternative allele. A dense common variant occupies about 0.24 MB for one million samples, against about 3.8 MB for the corresponding text VCF field. Each sparse carrier record is a pair of 32-bit integers: the top 5 bits index the ancestry (up to 32 ancestries) and the bottom 27 bits index the haplotype (up to $1.34 \times 10^8$ haplotypes, i.e. $6.7 \times 10^7$ individuals). The dense-to-sparse storage cost ratio is approximately 64:1, so a variant is stored sparsely below and densely above an alternative allele frequency of $1/64 \sim 0.0156$. This hybrid representation reduces storage and matches the two access patterns used downstream, dense marker streaming and per-variant retrieval, both used by FELIXassoc.

##### 1.3 Chunked streaming input and output

FELIXla is decoded in C++ in marker chunks ($\sim$ 1000 markers) and the on-disk blocks are memory-mapped rather than read whole, so the resident set is bounded by one chunk of dosages plus the downstream accumulators rather than by a full chromosome. Condition factors used by FELIXassoc are cached per ancestry block. This single-pass streaming design is what allows FELIXassoc to access ancestry-resolved dosages at biobank scale.

#### Supplementary Note 2. FELIXassoc association tests

##### 2.1 Score tests under the shared-effect and ancestry-specific models

FELIXassoc fits one null generalized linear mixed model per phenotype (Online Methods)[4,5] and reuses it for all variants. Testing ancestry-resolved dosages extends the local-ancestry association framework of Tractor[9] to a full mixed-model score test[4,8]. The cohort is modeled as an admixture of $K$ ancestries. For individual $i$ at variant $m$, let $G_{im}^{(a)}$ be the ancestry-$a$ alternative allele dosage and $L_{im}^{(a)} \in \{0,1,2\}$ the ancestry-$a$ local-ancestry haplotype count, with $\sum_{a=1}^{K} L_{im}^{(a)} = 2$. Under the ancestry-specific (heterogeneous) model each background carries its own per-allele effect and local ancestry enters as a covariate,

$$g(\mu_i) = \mathbf{C}_i^{\mathsf{T}} \boldsymbol{\alpha} + \sum_{a=1}^{K} \beta_a \, G_{im}^{(a)} + \sum_{a=1}^{K-1} \eta_a \, L_{im}^{(a)} + b_i, \qquad H_0 : \beta_1 = \cdots = \beta_K = 0,$$

where $\mathbf{C}_i$ is the covariate vector with fixed effects $\boldsymbol{\alpha}$, $\beta_a$ is the per-allele effect on the ancestry-$a$ background, $\eta_a$ is the effect of the ancestry-$a$ haplotype count, $b_i$ is the polygenic random effect, and $\mu_i = \mathbb{E}(Y_i)$. Only $K-1$ haplotype-count terms appear because the counts sum to a constant ($\sum_k L_{im}^{(a)} = 2$), so the $K$-th is redundant with the intercept; these terms adjust for admixture linkage disequilibrium and are included adaptively (Supplementary Note 2.5). The score test targets all $K$ per-allele effects $\boldsymbol{\beta}$; unlike the haplotype counts, the ancestry-specific dosages $G_{im}^{(a)}$ are not sum-constrained, so all ancestry-specific signals are identifiable and the heterogeneous test carries $K$ degrees of freedom.

Let $\mathbf{Y} = (Y_1, \ldots, Y_N)^{\mathsf{T}}$ be the phenotype vector, $\hat{\boldsymbol{\mu}}$ the fitted null mean, $\mathbf{G}_m$ the $N \times K$ matrix whose columns hold the ancestry-specific dosages, and $\tilde{\mathbf{G}}_m = \mathbf{G}_m - \mathbf{C}(\mathbf{C}^{\mathsf{T}}\mathbf{W}\mathbf{C})^{-1}\mathbf{C}^{\mathsf{T}}\mathbf{W}\mathbf{G}_m$ the covariate-adjusted dosage matrix ($\mathbf{C}$ the covariate matrix). The $K$-vector score and the heterogeneous statistic are

$$\mathbf{U}_m = \tilde{\mathbf{G}}_m^{\mathsf{T}}(\mathbf{Y} - \hat{\boldsymbol{\mu}}), \qquad T_{\text{het},m} = \mathbf{U}_m^{\mathsf{T}} \, \mathbf{V}_m^{-1} \, \mathbf{U}_m, \tag{S2}$$

where $\mathbf{U}_m$ is the difference between observed and null-expected ancestry-specific allele counts and $\mathbf{V}_m$ is its $K \times K$ null covariance. $T_{\text{het},m}$ is compared with a chi-squared distribution on rank($\mathbf{V}_m$) degrees of freedom (the number of tested ancestries). The GRM-free working covariance is

$$\mathbf{V}_m^0 = \tilde{\mathbf{G}}_m^{\mathsf{T}} \mathbf{W} \tilde{\mathbf{G}}_m, \tag{S3}$$

with $\mathbf{C}$ the covariate matrix and $\mathbf{W}$ the diagonal working-weight matrix from the fitted null model ($W_{ii} = \hat{\mu}_i(1-\hat{\mu}_i)$ for binary traits, and $\mathbf{W} = \phi^{-1}\mathbf{I}$ with $\phi$ the residual variance for quantitative traits). The score covariance used in (S2) is the variance-ratio-adjusted $\mathbf{V}_m = \hat{r}\,\mathbf{V}_m^0$ (Supplementary Note 2.2).

Under the shared-effect model, $\mathbf{G}_m$ is replaced by the aggregate dosage $G_{im} = \sum_k G_{im}^{(a)}$, and $H_0 : \beta_G = 0$ is tested with the one-degree-of-freedom form of (S2), which reduces to the standard single-variant score test.

##### 2.2 Variance-ratio approximation

The exact score variance under the fitted mixed model is $\mathbf{V}_m^{\text{full}} = \mathbf{G}_m^{\mathsf{T}} \mathbf{P} \, \mathbf{G}_m$, with the null-model projection $\mathbf{P}$ and working covariance $\boldsymbol{\Sigma} = \mathbf{W}^{-1} + \tau \boldsymbol{\Psi}$ ($\boldsymbol{\Psi}$ the sparse GRM, $\tau$ the polygenic variance) defined in the Online Methods and estimated by penalized quasi-likelihood[6] with average-information REML[7]. Forming $\mathbf{P}$, an $N \times N$ matrix, for every variant is infeasible at biobank scale. As in SAIGE[4] and related mixed-model methods[10,11,12], FELIXassoc uses the empirical observation that the ratio of $\mathbf{V}_m^{\text{full}}$ to its covariate-projected, GRM-free counterpart $\mathbf{V}_m^0$ (S3) is approximately constant across variants,

$$\hat{r} = \frac{\mathbf{V}_m^{\text{full}}}{\mathbf{V}_m^0}. \tag{S4}$$

The ratio $\hat{r}$ is estimated from a random subset of variants and applied genome-wide, so the score covariance for every variant is $\mathbf{V}_m = \hat{r}\,\mathbf{V}_m^0$, formed from covariate-adjusted dosages without repeated inversion of the fitted covariance.

##### 2.3 Saddlepoint adjustment for imbalanced binary traits

For binary traits with imbalanced case-control ratios the normal approximation to the score-statistic distribution inflates type I error, particularly for low-frequency variants. FELIXassoc applies the saddlepoint approximation (SPA)[13,14]. For the shared-effect test SPA is applied directly to the one-dimensional score statistic. For the ancestry-specific test SPA is applied to each marginal ancestry-$a$ score statistic, giving a per-ancestry ratio $c_k = \mathrm{Var}_{\mathrm{SPA},k} \,/\, \mathrm{Var}_{\mathrm{N},k}$ of the SPA-adjusted to normal-approximation variance. The joint score covariance is then rescaled elementwise,

$$\hat{V}_{kl} = \sqrt{c_k\, c_l}\; V_{kl}, \tag{S5}$$

before the multi-degree-of-freedom statistic (S2) is formed. As in SAIGE[4], SPA is applied only when the absolute normal-approximation $z$-score exceeds 2; nearer the null the normal approximation is used, where SPA is numerically unstable and offers little benefit.

##### 2.4 Cauchy combination of the two tests

Let $P_{\mathrm{hom}}$ and $P_{\mathrm{het}}$ be the shared-effect and ancestry-specific $P$ values. FELIXassoc combines them with the Cauchy combination test[15],

$$T_{\mathrm{CCT}} = \sum_{j\in\{\mathrm{hom},\mathrm{het}\}} w_j \tan\big[(0.5 - P_j)\pi\big], \qquad P_{\mathrm{CCT}} = 0.5 - \frac{\arctan(T_{\mathrm{CCT}})}{\pi}, \tag{S6}$$

with equal weights $w_{\mathrm{hom}} = w_{\mathrm{het}} = 0.5$. The Cauchy combination is valid under arbitrary dependence between the component statistics and requires no estimate of their correlation. The combined $P$ value is asymptotically equivalent to the minimum of the component $P$ values scaled by the number of tests, so power tracks the better-fitting model at each locus.

##### 2.5 Adaptive local-ancestry conditioning

Where the local ancestry haplotype count is strongly associated with both dosage and phenotype, marginal ancestry-specific tests can return mirrored effect estimates, because the dosage assigned to one ancestry is negatively correlated with the dosage or local ancestry of another (admixture linkage disequilibrium). At each variant, FELIXassoc first tests the association between the phenotype and each local ancestry haplotype count $L_{im}^{(a)}$ with a marginal score test, applying SPA for imbalanced binary traits. If a haplotype-count association reaches a configurable significance threshold ($P < 5 \times 10^{-6}$ in these analyses), the corresponding counts are treated as nuisance covariates in a conditional score test[8,16]. Let $\mathbf{U}_g$ and $\mathbf{U}_L$ be the genotype and selected haplotype-count score vectors, which jointly follow a multivariate normal distribution under the null with covariance

$$\begin{pmatrix} \mathbf{\Sigma}_{gg} & \mathbf{\Sigma}_{gL} \\ \mathbf{\Sigma}_{Lg} & \mathbf{\Sigma}_{LL} \end{pmatrix}. \tag{S7}$$

The conditional genotype score and its covariance are $\mathbf{U}_{g|L} = \mathbf{U}_g - \mathbf{\Sigma}_{gL}\mathbf{\Sigma}_{LL}^{-1}\mathbf{U}_L$ and $\mathbf{\Sigma}_{g|L} = \mathbf{\Sigma}_{gg} - \mathbf{\Sigma}_{gL}\mathbf{\Sigma}_{LL}^{-1}\mathbf{\Sigma}_{Lg}$, which separate the allelic association from the association attributable to local ancestry exposure. At loci where local ancestry carries little information about dosage the marginal test is reported, so an ancestry-specific variant that is too rare for ancestry to predict carrier status is not conditioned unnecessarily.

#### Supplementary References


1. Delaneau, O., Marchini, J. & Zagury, J.-F. A linear complexity phasing method for thousands of genomes. *Nat. Methods* **9**, 179–181 (2011).
2. Browning, S. R., Waples, R. K. & Browning, B. L. Fast, accurate local ancestry inference with FLARE. *Am. J. Hum. Genet.* **110**, 326–335 (2023).
3. Maples, B. K., Gravel, S., Kenny, E. E. & Bustamante, C. D. RFMix: a discriminative modeling approach for rapid and robust local-ancestry inference. *Am. J. Hum. Genet.* **93**, 278–288 (2013).
4. Zhou, W. et al. Efficiently controlling for case-control imbalance and sample relatedness in large-scale genetic association studies. *Nat. Genet.* **50**, 1335–1341 (2018).
5. Lin, X., Dey, R., Li, X. & Li, Z. Scalable analysis of large multi-ancestry biobanks by leveraging sparse ancestry-adjusted sample-relatedness. *Res. Sq.* (2024) doi:10.21203/rs.3.rs-5343361/v1.
6. Breslow, N. E. & Clayton, D. G. Approximate inference in generalized linear mixed models. *J. Am. Stat. Assoc.* **88**, 9–25 (1993).
7. Gilmour, A. R., Thompson, R. & Cullis, B. R. Average information REML: an efficient algorithm for variance parameter estimation in linear mixed models. *Biometrics* **51**, 1440–1450 (1995).
8. Chen, H. et al. Control for population structure and relatedness for binary traits in genetic association studies via logistic mixed models. *Am. J. Hum. Genet.* **98**, 653–666 (2016).
9. Atkinson, E. G. et al. Tractor uses local ancestry to enable the inclusion of admixed individuals in GWAS and to boost power. *Nat. Genet.* **53**, 195–204 (2021).
10. Svishcheva, G. R. et al. Rapid variance components-based method for whole-genome association analysis. *Nat. Genet.* **44**, 1166–1170 (2012).
11. Loh, P.-R. et al. Efficient Bayesian mixed-model analysis increases association power in large cohorts. *Nat. Genet.* **47**, 284–290 (2015).
12. Jiang, L. et al. A resource-efficient tool for mixed model association analysis of large-scale data. *Nat. Genet.* **51**, 1749–1755 (2019).
13. Daniels, H. E. Saddlepoint approximations in statistics. *Ann. Math. Stat.* **25**, 631–650 (1954).
14. Dey, R., Schmidt, E. M., Abecasis, G. R. & Lee, S. A fast and accurate algorithm to test for binary phenotypes and its application to PheWAS. *Am. J. Hum. Genet.* **101**, 37–49 (2017).
15. Liu, Y. & Xie, J. Cauchy combination test: a powerful test with analytic p-value calculation under arbitrary dependency structures. *J. Am. Stat. Assoc.* **115**, 393–402 (2020).
16. Yang, J. et al. Conditional and joint multiple-SNP analysis of GWAS summary statistics identifies additional variants influencing complex traits. *Nat. Genet.* **44**, 369–375 (2012).
